# Abundance of *Bifidobacterium* species in the infant gut microbiota and associations with maternal-infant characteristics in Dhaka, Bangladesh

**DOI:** 10.1101/2025.03.03.25323106

**Authors:** Aline C. Freitas, Grace Li, Jakaria Shawon, Huma Qamar, Lisa G. Pell, Mamun Kabir, Ovokeraye H. Oduaran, Scarlett Puebla-Barragan, Diego G. Bassani, Karen M. O’Callaghan, Jennifer C. Onuora, Miranda G. Loutet, Cole Heasley, Cody W. E. Starke, Abdullah Al Mahmud, Davidson H. Hamer, Eleanor Pullenayegum, Md Iqbal Hossain, Md Muniruzzaman Siddiqui, Mohammad Shahidul Islam, Philip M. Sherman, Prakesh S. Shah, S. M. Abdul Gaffar, Shamima Sultana, Shaun K. Morris, Tahmeed Ahmed, Rashidul Haque, Shafiqul Alam Sarker, Daniel E. Roth

## Abstract

The early infant gut microbiota is dominated by bifidobacteria, but there is substantial variation at the (sub)species level. Patterns of postnatal *Bifidobacterium* subspecies colonization in low or middle-income countries have not been widely studied. We used (sub)species-specific qPCR to quantify *B. infantis, B. longum*, and *B. breve* in stool samples from 1132 infants (0-6 months) in urban Dhaka, Bangladesh. *B. infantis* absolute abundance started low but increased in the first two months, whereas *B. longum* and *B. breve* abundances remained comparatively low. *B. infantis* emerged earlier in infants delivered by C-section, but by ∼2 months of age, infants delivered by C-section or vaginally had similar *B. infantis* abundances. Infant antibiotic exposure, feeding patterns, and maternal stool *B. infantis* were not associated with infant *B. infantis*. In settings where *B. infantis* is widespread, its patterns of postnatal colonization can be used to inform the design of targeted microbiota-modifying interventions in infancy.

## INTRODUCTION

Imbalances in the infant gut microbiota have been associated with adverse health outcomes, such as necrotizing enterocolitis, sepsis, and acute malnutrition^1^. In infants born at term, the gut microbiota is generally colonized by bifidobacteria, a biomarker of health in early infancy^2^. However, patterns in *Bifidobacterium* species colonization and the timing of emergence are inconsistent across populations, underscoring the need for longitudinal studies spanning diverse geographical settings. *Bifidobacterium longum* subspecies *infantis* (*B. infantis*) in particular has been the focus of considerable interest given its role in enhancing gut barrier function, promoting acidification of the intestinal lumen via production of short chain fatty acids, and association with enhanced immune responses to infant vaccination^3,4,5^. *B. infantis* is believed to be the only bacterium capable of digesting all human milk oligosaccharides (HMOs), which confers a selective competitive advantage against other bacteria^6^. Nevertheless, few microbiome-related studies have examined early postnatal colonization patterns at the subspecies level in infants in low- and middle-income countries (LMIC).

In a longitudinal cohort study of infants residing in an urban area in Dhaka, Bangladesh, we used (sub)species-specific quantitative real-time PCR (qPCR) to describe the timing of emergence and absolute abundance (AA) trajectory of three *Bifidobacterium* targets in infant stool samples from birth up to six months of age: *Bifidobacterium longum* subspecies *infantis* (hereafter *B. infantis*), *Bifidobacterium longum* subspecies *longum* (hereafter *B. longum*) and *Bifidobacterium breve* (*B. breve*). Using multivariable quantile regression models, we assessed whether infant-maternal characteristics (including mode of delivery, infant feeding practices, infant antibiotic exposure, and maternal stool *B. infantis* detection) were associated with *B. infantis* abundance. We also investigated the dynamics between *B. infantis* and *B. longum/B. breve* AA patterns as synergistic or competitor (sub)species and used metagenomics in a subset of samples to confirm that *B. infantis* was abundant and dominant in this population. This study provides detailed descriptions of the natural patterns of *Bifidobacterium* species colonization in an urban infant population residing in Bangladesh, a lower middle-income country in South Asia, a region that has been historically underrepresented in microbiome-related studies^7^. The new insights into the timing and patterns of postnatal colonization can be used to inform the design of targeted microbiota-modifying interventions in infancy.

## MATERIALS AND METHODS

### Study design and participants

This is a longitudinal prospective cohort study based on the SEPSiS (Synbiotics for the Early Prevention of Severe Infections in Infants) Observational cohort. Ethical approval was provided by the Research Ethics Board (The Hospital for Sick Children; no.: 1000063899, and the Ethical Review Committee (International Centre for Diarrhoeal Disease Research, Bangladesh, icddr,b; no. PR-19045); The study was registered at clinicaltrials.gov (NCT04012190). Enrollment was conducted at two public hospitals in Dhaka, Bangladesh: the Maternal and Child Health Training Institute (MCHTI) and the Mohammadpur Fertility Services and Training Centre (MFSTC), between November 2020 and October 2021. The Director General of Family Planning (DGFP), Government of Bangladesh approved the implementation of study activities at MCHTI and MFSTC.

To be eligible for enrolment into the Observational cohort study, infants needed to: be 0-4 days old (day of birth was defined as day 0); delivered at a study hospital; orally feeding at the time of screening; and their caregivers planned to maintain residence within the study catchment area until 60 days of age. Infants were not eligible to be enrolled if: they had a birth weight <1500g; death or major surgery was considered highly probable within first week of life; they had a major congenital anomaly of the gastrointestinal tract; there was evidence of maternal HIV infection and/or history of receiving anti-retroviral drugs for presumed HIV infection; infants were receiving mechanical ventilation and/or cardiac support; parenteral antibiotics at the time of recruitment (allowed to be enrolled if antibiotics were stopped before recruitment day, up to day 4); prenatal or postpartum use of non-dietary probiotic supplements by the mother; administration of non-dietary probiotic or prebiotic supplements to the infant; enrollment of infant in another clinical trial involving the administration of probiotics and/or prebiotics; resides in the same household as another infant previously enrolled in any study within the SEPSiS platform who is currently <60 days of ag e (twins may be enrolled simultaneously); multiple gestation with three or more liveborn infants. Additional eligibility criteria for inclusion in the present study included: at least one infant stool sample available with valid qPCR results (i.e., passed qPCR QC, described in the methods). Written informed consent was obtained from the parent/guardian.

### Data and sample collection

Clinical data were collected by trained study personnel at baseline (day of enrolment, 0-4 days) and at routine clinical visits (3 and 6 days after baseline, and at 10, 14, 21, 28, 35, 60, 90, 180 days postnatal age), during in-person home or hospital visits or by telephone when an in-person visit was not feasible. Infant stool samples were collected according to a schedule (A, B, or C) assigned at enrollment. Infants in schedules A and B were enrolled at 0-1 day of age and had up to 11 stool samples collected (0-1, 3-4, 6-7, 10, 14, 21, 28, 35, 60, 90, 180 days postnatal age). For schedule A, sampling was stratified by mode of delivery to ensure that an approximately equal number of infants were delivered vaginally and via C-section. Infants in schedule C were enrolled at 0-4 days of age, with a maximum of 3 stool samples collected (day of enrolment plus two additional samples up to day 60, timing randomized). Schedule A was primarily designed for the purpose of metagenomic analyses and schedule B was designed primarily for the purpose of detailed longitudinal qPCR analysis of *B. infantis*; however, all participants enrolled in the SEPSiS Observational Study, irrespective of collection schedule, served as the pool from which samples for this study were selected.

Infant stool samples from schedules A and B were collected, homogenized, and aliquoted by a trained study worker and then frozen in the vapor phase of liquid nitrogen within 20 minutes of defecation (tier 1).

For schedule C, 46% of samples were collected using the tier 1 protocol, and 54% of samples were collected by the caregiver and placed in a cold box within 20 minutes of defecation and then homogenized, aliquoted and frozen by a study worker within 6 hours of defecation (tier 2). Maternal samples were collected during the first week postpartum in a manner that was similar to the infant tier 2 protocol. All stool specimens were stored at −70°C or colder until ready for analysis.

### DNA extraction for qPCR

Total DNA was extracted from 100-150 mg of stool using mechanical disruption by bead beating and the QIAamp Fast DNA Stool Mini Kit (QIAGEN) following the manufacturer’s protocol. Total DNA was eluted in 200 μL. DNA extraction was performed in batches of up to 47 samples with one extraction blank (EB). DNA was quantified using a Nanodrop 2000 spectrophotometer (Thermo Scientific) and samples with a DNA concentration below the limit of detection (LOD) of the Nanodrop (i.e., <2 ng/μL) were assigned an imputed value of 1 ng/μL. DNA extraction and qPCR were performed in the Parasitology Laboratory, at the icddr,b.

### qPCR assay

Individual TaqMan-based (sub)species-specific quantitative PCR (qPCR) assays were used to quantify *B. infantis* (subspecies), *B. longum* (subspecies) and *B. breve* (species) in stool samples using primers and probes previously reported^8,9^ (**Table S1**).

qPCR was performed in 96-well plates, with each plate including duplicates of no-template control (NTC), EB and standards; unknown samples were run in single wells. Phocid herpes virus (PhHV) ( European Virus Archive – Global) was used as an internal control in the *B. infantis* qPCR (i.e., duplex assay) in which 10 μL of 10X PhHV was added to each stool sample prior to DNA extraction; the *B. longum* and *B. breve* qPCR were each run as single-plex assays. For all targets, each reaction contained 1x iTaq Universal Probes Supermix (Bio-Rad), 1 μM of each primer (Forward (F), and Reverse (R)), 250 nM probe, and 5 μL of template DNA or water (NTC); final volume was 20 μL. The *B. infantis* assay also had 1 μM of PhHV each primer (F, R) and 250 nM probe. The reaction was performed on a CFX96™ Real-Time System (Bio-Rad) and consisted of a 5 min denaturation step at 95°C, followed by 40 cycles of 15 s at 95°C and 1 min at 61°C.

Standard curves were generated using genomic DNA extracted with QIAamp fast DNA stool mini kit (QIAGEN) from pure cultures of *B. longum* subsp. *infantis* ATCC 15697, *B. longum* subsp. *longum* ATCC 15707 and *B. breve* ATCC 15700. Prior to DNA extraction, cells from a liquid culture were counted using hemocytometry; enumerated cell suspensions were centrifuged at 10,000 × *g* for 5 minutes, and the cell pellets were stored at −80°C until DNA was extracted.

The DNA extract from each cell culture was 10-fold diluted, and the upper and lower limit of quantification (ULOQ and LLOQ) were estimated based on the most concentrated and the 10^5^-fold diluted standard, respectively. Standards were generated from more than one cell culture, thus, the quantifiable range of the assay differed based on which cell culture was used: 9.26 x 10^1^ to 9.26 x 10^6^ or 1.22 x 10^1^ to 1.22 x 10^6^ cells for *B. infantis,* 3.2 x 10^2^ to 3.2 x 10^7^ or 1.32 x 10^2^ to 1.32 x 10^7^ cells for *B. longum,* and 3.2 x 10^2^ to 3.2 x 10^7^ cells for *B. breve.* Samples with starting quantities above the ULOQ were extrapolated from the standard curve after experimentally confirming that 100-fold diluted repeats had similar cell counts as the undiluted sample calculated by extrapolating the standard curve.

The LOD for *B. infantis* was determined as the starting quantity corresponding to a Cq value 35 for each qPCR plate (i.e., sample considered below LOD if Cq ≥35), with a range across plates of 4 to 50 cells. The threshold Cq value of 35 coincided with the minimum Cq value used to identify potential contamination of EBs. The LOD for *B. longum* and *B. breve* was determined experimentally to be 1 and 3 cells per reaction, respectively, after confirming the linearity of the standard curve between the LLOQ and the LOD. For all targets, samples with starting quantities between the LLOQ and the LOD were extrapolated from the standard curve.

Results for qPCR were reported as Absolute Abundance (AA), normalized to total extracted DNA mass and expressed as cells per microgram of DNA (cells per μg DNA). AA normalized to stool mass (cells per gram of stool) and not normalized (cells) were used in sensitivity analyses. Samples that did not amplify or had starting quantities below the LOD were imputed with a single value corresponding to half of the LOD (in cells) and then normalized to the sample’s DNA mass to generate ‘cells per μg DNA’. The median AA normalized to DNA mass among these samples was subsequently used as a single imputation value in analyses; the imputed value was calculated separately for each target. AA was log _10_-transformed prior to analysis.

### qPCR Quality Control

For all targets, only qPCR plates that passed all of the following quality control (QC) criteria were included in analyses: NTC Cq >38; R^2^ from the standard curve >0.96; efficiency between 89.5% and 110%; coefficient of variation (CV) between standard replicates (intra-assay) <10%; EB with median Cq <35 or one replicate Cq <33. DNA extraction was repeated using a new stool aliquot for samples that had DNA extracted on the same day as an EB that failed QC criteria. The empirical range of the inter -assay CV for the standards Cq was 2.1-3.9% (*B. infantis*), 2.1-5.2% (*B. longum*), and 2.4-5.5% (*B. breve*).

Inter-assay concordance for unknown samples was assessed by repeating the *B. infantis* assay for a subset of samples (n=62) using the same DNA extract. Of the 62 samples, 43 were at or above LOD in both assays, with a CV range of 0.1-5.2% (based on the log cells) and an Intraclass Correlation Coefficient (ICC) of 0.997 (95% CI: 0.916 - 0.999); the other 19 samples were below LOD in both assays (100% agreement between runs).

Lastly, a spike-in test was conducted in which known numbers of *B. infantis* cells (2.45 x 10^6^, 2.45 x 10^7^, 2.45 x 10^8^) were added to each of five stool samples prior to DNA extraction (3 x 5, n=15 tests). A parallel DNA extraction was conducted for the same set of samples without spike-in where 2/5 samples were below LOD, and 3/5 samples had 5.2, 5.7 and 8.3 log cells per ug DNA. The range of recovery was 91-100% (median 96%), which was defined as the percentage of observed cell number (as log cells per μg DNA) in relation to the expected cell number (where the expected number was the sum of cells in the non-spiked sample plus the number of added cells from the spike-in).

### qPCR verification of false negatives of test results

Given the large proportion of early postnatal samples that contained undetectable levels of *B. infantis*, four different analyses were conducted to confirm that samples that did not amplify in the qPCR assay were not false negatives. First, among samples that were tested for all three bacterial targets, PhHV, which was included in the duplex qPCR assay as an internal control, amplified in 99.5% (1114/1120) of samples; despite non-amplification of PhHV in a small proportion of samples, there was no difference in the probability of detecting *B. infantis, B. longum* or *B. breve* based on PhHV amplification status (Fisher’s exact test, all p >0.5). Second, *B. infantis* qPCR was repeated using a different stool aliquot from the same stool sample for a subset of samples that did not amplify for any target; 96% (81/84) of samples did not amplify in the repeat run indicating that differences in stool aliquot batches were not a main source of variation that could lead to false negatives. Third, to determine whether non-amplification was due to saturation of the qPCR reaction (e.g., high DNA concentration or other inhibitors), we repeated the *B. infantis* qPCR assay at 5x, 10x, and 100x dilutions for 56 samples that did not amplify (56 x 3 dilutions = 168 tests); non-amplification persisted in most samples after dilution (148/168, 88%), indicating that non-amplification was unlikely due to inhibitors (among those that amplified, only 4/20 were above LOD, with a Cq range of 32.4-34.4; no sample amplified for all three dilutions). Finally, we also compared sample DNA concentration with qPCR detection status of each target to test the hypothesis that non-detection was due to low DNA concentration; detectable and non-detectable samples had a similar distribution of DNA concentrations, with no evidence that low DNA concentration was associated with non-detection status (**Figure S1**).

### Metagenomic shotgun sequencing and taxonomic profiling

A subset of infant stool samples from schedule A (n=55 infants, 357 samples) for which there were *B. infantis* qPCR data (i.e., aliquots from the same stool sample) was also analyzed using metagenomic shotgun sequencing. Total DNA was extracted from 15-40 mg of stool using MagMAX™ Microbiome Ultra Nucleic Acid Isolation Kit (Applied Biosystems), automated with the KingFisher Flex system (Thermo Fisher Scientific). DNA was quantified using the Quant-iT™ PicoGreen™ dsDNA Assay Kits (Invitrogen, Thermo Fisher Scientific) and normalized to 5 ng of DNA. The DNA library was constructed using KAPA HyperPlus Kit (Roche) with iTru adapters. Libraries were first submitted to shallow sequencing on the Illumina iSeq 100 system to better inform pooling volumes for a read count-based normalization^10^. Then, libraries were sequenced in a single lane on the Illumina NovaSeq 6000 s4 flow cell system, with a length of 150-bp paired-end reads. DNA extraction and library preparation were performed at the Knight Lab, UCSD (University of California San Diego) and sequencing was done at the Institute for Genomic Medicine, UCSD.

Reads were processed following the default workflow on the Qiita platform^11^ where reads were demultiplexed, quality filtered, and adapters trimmed using *fastp*^12^. Internal control PhiX was removed using *Minimap2*^13^. Reads were then filtered against the human genomes GRCh38 and T2T-CHM13v2.0 using *Minimap2*, and against the pangenome from the Human Pangenome Reference Consortium^14^ using *Movi*^15^. Samples with fewer than 500,000 quality filtered reads were excluded. An average of 5,218,284 reads per sample (n=332) were used for downstream analysis. Taxonomic profiling was done using MetaPhlAn 4^16^ with a custom database tailored for *B. longum* subspecies identification (standard database mpa_vJun23_CHOCOPhlAnSGB_202403 with *B. longum* subspecies specific markers), as previously described^17^. Results were reported as Relative Abundance (RA).

## STATISTICAL ANALYSIS

Quantile regression analyses were performed in Stata/BE 18.0 with the *qreg2* module^18^. Other statistical analyses and graphical representations were performed in R (version 4.4.1). Continuous data were expressed as median and interquartile range (IQR). The number of participants or samples included in each analysis is annotated in the figures and tables.

### Variable definitions

Infant antibiotic exposure was defined at the sample level as any exposure to systemic (i.e., administered orally or via injection) antibiotics from birth until the collection of a stool sample (exposed vs. never exposed). Antibiotic use data were collected from both inpatient and outpatient settings. Human milk feeding pattern was based on the least stringent feeding pattern from birth until stool collection, and was defined as (from most to least stringent): exclusive (no additional food, water, or other fluids, except for medicines, vitamins, or mineral drops), predominant (human milk in addition to water, sugar water, honey, or other non-milk, non-formula liquids), partial (human milk in addition to formula, milk-based products, any other liquids, solid or semi-solid foods), or, none (no human milk consumption at least once up to the time of stool sample collection).

### Outcomes and summary measures

The primary outcome was absolute abundance (AA) (cells per μg DNA), as a continuous variable. Secondary outcomes were 1) binary variable based on whether the target was detected (at or above LOD) or not (below LOD) by qPCR; 2) binary ‘high’ or ‘low’ abundance category, whereby the midpoint between the AA peaks of the bimodal distribution of samples was used as the threshold for dichotomization: samples with cell counts above the threshold were categorized as ‘high’ AA samples, and samples with cell counts below the threshold were categorized as ‘low’ AA samples. Median AA (further described below) and prevalence (defined as the proportion of samples in which the bacterial target was detected) were used as group-level summary measures.

### Primary analysis: B. infantis, B. longum and B. breve AA over infant age

A regression model with clustered standard errors and restricted cubic splines (knots at 7, 14, 28, and 60 days of age) was used to estimate the median AA with 95% confidence intervals (95% CI) of *B. infantis*, *B. longum* and *B. breve* in infant stool samples during the first six months since delivery. AA was treated as a dependent continuous variable, and age was the primary independent continuous variable.

### Primary analysis: B. infantis AA, by infant-maternal characteristics

Models were extended to assess the effect of different exposures (mode of delivery, infant exposure to antibiotics and infant feeding pattern) on *B. infantis* AA. In each multivariable model, the exposure was included as a main effect and in an interaction term with age. The models were adjusted for putative confounders based on their clinical relevance, identified by Directed Acyclic Graphs (DAG) (Mode of delivery: enrollment hospital, gestational age, maternal age, parity, maternal education, asset index. Infant antibiotics: enrollment hospital, gestational age, mode of delivery, maternal education, asset index. Feeding pattern: enrollment hospital, gestational age, mode of delivery, length of postnatal hospital stay, maternal age, parity, maternal education, asset index, household size) (**Figure S2**). Confounding was examined based on stratified analysis by potential confounders. Predicted medians and 95% CI at specific ages of interest and between-group differences in medians were estimated as marginal estimates based on the models described above.

DAGs were also used to identify potential mediators that could explain the association between the exposure of interest and *B. infantis* AA. Mediation analysis was conducted when the candidate mediator was both associated with the exposure and the outcome^19^; candidate mediators were included in the model (one at a time), restricted to age windows of interest, and attenuation of the coefficient for the exposure of interest was interpreted as evidence of possible mediation.

As a sensitivity analysis, these multivariable models were restricted to participants in schedules A and B, who, by study design, provided more stool samples spanning the first six months of postnatal age in comparison with schedule C infants who provided fewer samples (maximum 4, median: 2 samples) that were collected mostly up to two months of age. This sensitivity analysis was also intended to examine possible effects of the COVID-19 pandemic and calendar date on *B. infantis* AA given that schedules A and B recruitment occurred over a shorter timeframe (May-Oct 2021) and after the launch of schedule C recruitment (Nov 2020-Oct 2021).

### AA relationships between targets

Differences in the proportion of samples with a low vs. high AA of *B. infantis* among samples with low vs. high AA of *B. longum* or *B. breve* was compared within a sample using a Chi-square test in R (*ggstatsplot*^20^). Differences were considered statistically significant when p < 0.05.

### Probability of samples having a high AA of B. infantis, by infant-maternal characteristics

Binary AA of *B. infantis* was modeled as a function of age using Generalized Estimating Equations (GEE) with a logit link, binomial distribution, robust standard errors, and restricted cubic splines (knots at 7, 14, 28, and 60 days of age), adjusted for confounders as described above. The predicted probabilities of samples belonging to high AA category was determined and plotted over infant age with 95% CIs derived from the predicted probabilities and used to compare the effect of exposures within six months of age. Analyses were conducted in R with *geepack*^21^ and *marginaleffects*^22^.

### Association between detection of B. infantis in maternal stool and AA of B. infantis in newborn stool samples

The association between maternal stool *B. infantis* (detected vs. not detected) within the first week postpartum and newborn *B. infantis* AA within the first month of age (3-31 days) was assessed using quantile regression. Coefficients of unadjusted and adjusted models (by study site and mode of delivery) were examined. Maternal *B. infantis* was treated as a time-fixed, independent binary (detected vs. not-detected) variable derived using qPCR data from the first maternal stool sample available within 0-7 days postpartum. Infant samples that were collected prior to the maternal sample were excluded from the analysis.

Chi-square test was used to investigate the association of maternal stool *B. infantis* detection (binary variable) and socioeconomic factors: asset index (categorical: 1-5) and maternal education (categorical: little to no schooling, secondary incomplete, secondary complete or higher); analysis was restricted to maternal stool samples collected within the first week postpartum, as described above.

## RESULTS

### Participant characteristics

A total of 1132 infants were included in the present study (**Figure 1**). Infant, maternal, and household characteristics of participants are summarized in **Table 1**. In total, 60% of participants were enrolled at the Mohammadpur Fertility Services and Training Centre (MFSTC), and 40% at the Maternal and Child Health Training Institute (MCHTI). Most infants were born at term (92%), and just over half were delivered by C-section (54%). Approximately half (52%) of all infants were exclusively breastfed up to two months, and most (96%) were at least partially breastfed up to approximately six months of age. Most infants (62%) received either oral or parenteral antibiotics at least once within the first six months of age, and nearly all mothers (95%) were administered at least one dose of peripartum antibiotics. Differences in characteristics between participants enrolled at the two study hospitals are shown in **Table S2**.

**Figure 1.**
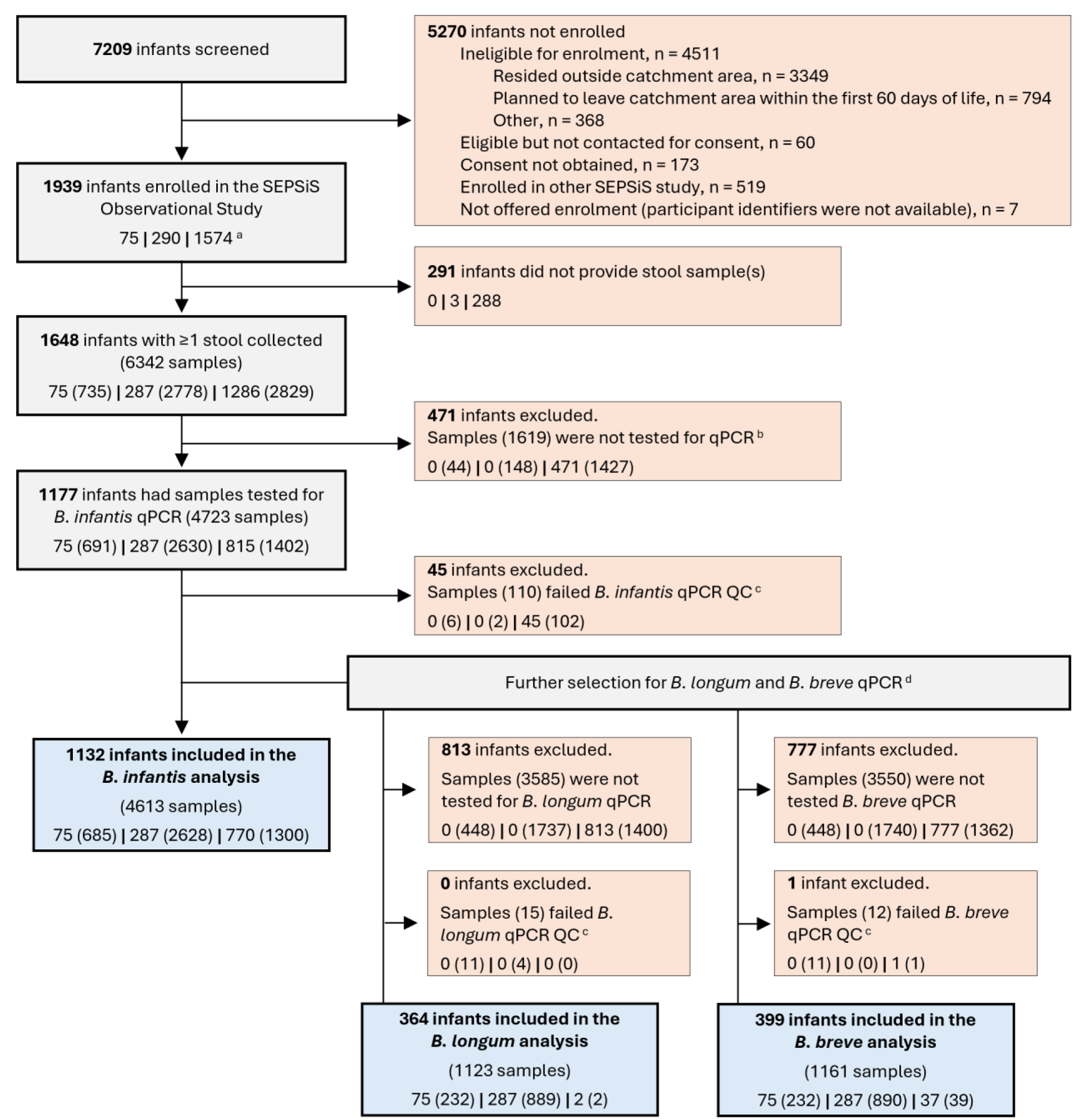
Study flow diagram. Participant enrollment and sample selection for qPCR. **^a^** Number of infants and samples (if applicable) are shown stratified by schedule, reported as: # infants (# samples) for schedules A | B | C, respectively. All numbers in parenthesis refer to samples. ^b^ Samples from schedules A and B were selected for *B. infantis* qPCR by the probability-based algorithm: a fixed selection probability of ∼0.67 (6/9) for 9 collected samples was applied to account for the likelihood of the collection of <11 expected samples per infant; first (earliest) sample was always included. Schedule C stool samples were only included in the qPCR prior to the launch of schedules A and B, with no selection criteria. ^c^ Samples were excluded from the study if they failed qPCR QC (quality control) and were not tested again, or if QC failure persisted after qPCR repeats. The criteria for passing QC are described in the Methods. ^d^ All schedules A and B samples analyzed for *B. infantis* served as the pool from which samples for *B. longum* and *B. breve* qPCR were selected (1-4 samples, random timing). Some schedule C samples were selected early in the study prior to the launch of schedules A and B.

**Table 1.**
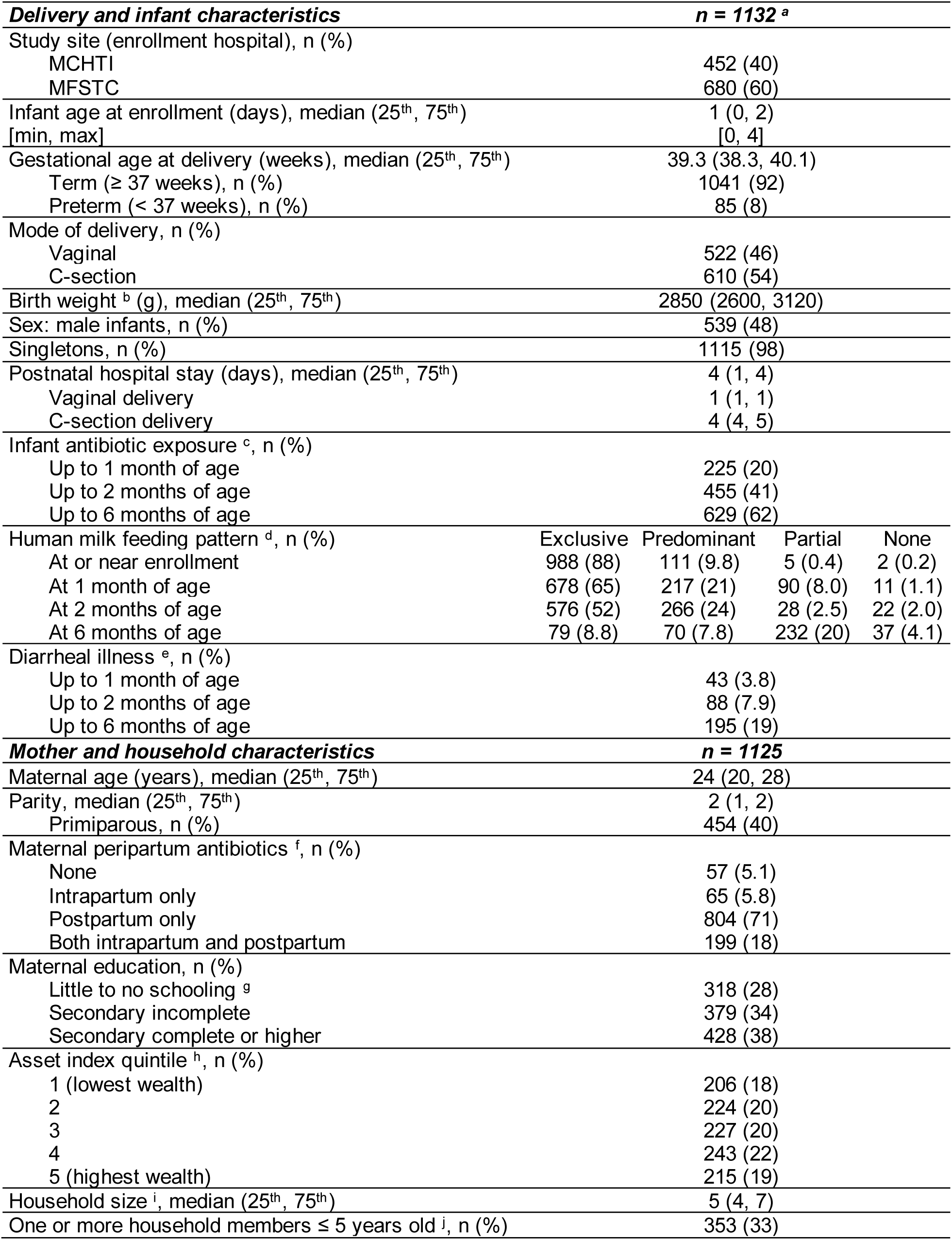
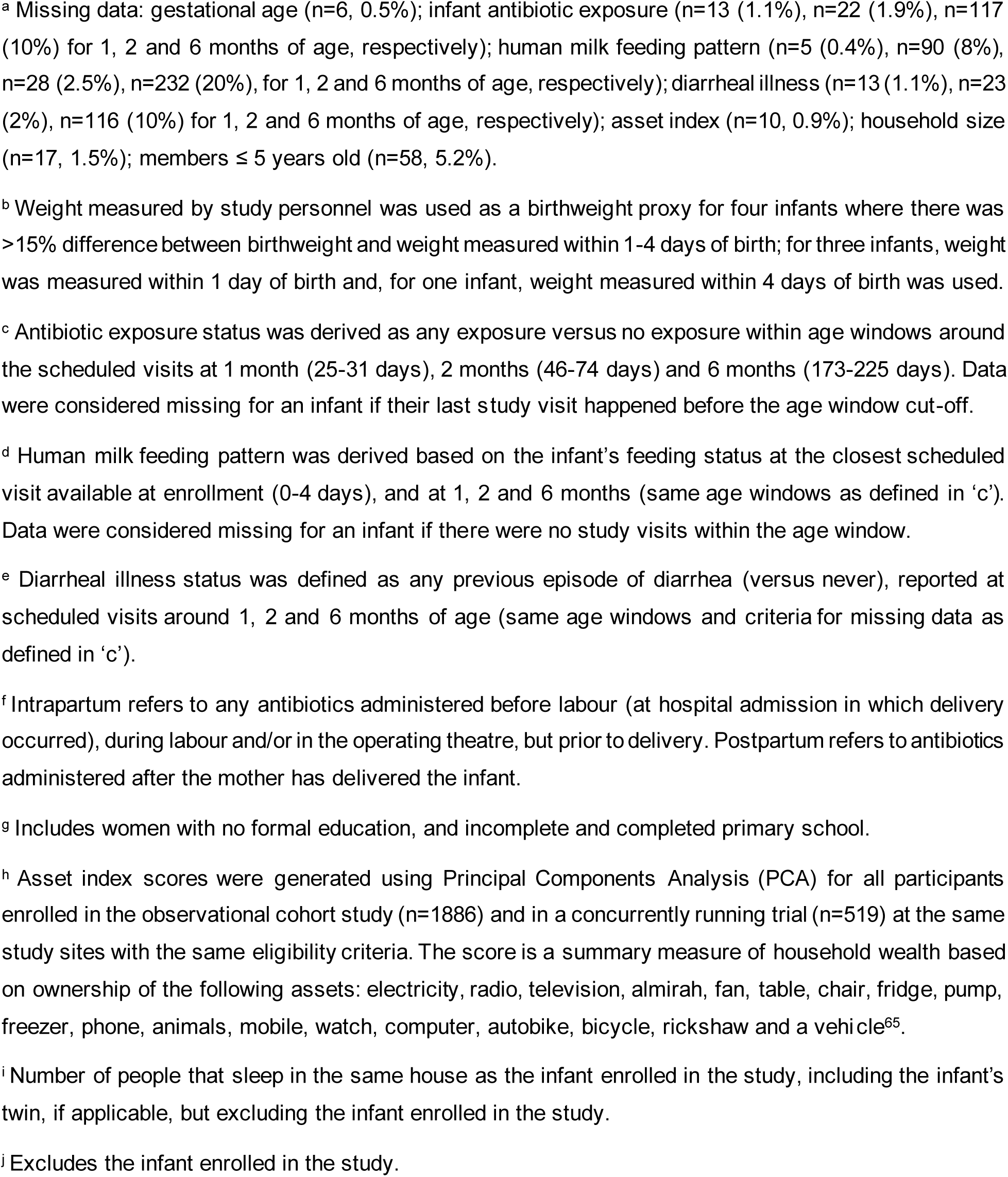
Infant, maternal, and household characteristics.

| ***Delivery and infant characteristics*** |  | ***n = 1132<sup>a</sup>*** |  |  |  |
| --- | --- | --- | --- | --- | --- |
| Study site (enrollment hospital), n (%) |  |  |  |  |  |
| MCHTI |  | 452 (40) |  |  |  |
| MFSTC |  | 680 (60) |  |  |  |
| Infant age at enrollment (days), median (25<sup>th</sup>, 75<sup>th</sup>) |  | 1 (0, 2) |  |  |  |
| [min, max] |  | [0, 4] |  |  |  |
| Gestational age at delivery (weeks), median (25<sup>th</sup>, 75<sup>th</sup>) |  | 39.3 (38.3, 40.1) |  |  |  |
| Term (≥ 37 weeks), n (%) |  | 1041 (92) |  |  |  |
| Preterm (< 37 weeks), n (%) |  | 85 (8) |  |  |  |
| Mode of delivery, n (%) |  |  |  |  |  |
| Vaginal |  | 522 (46) |  |  |  |
| C-section |  | 610 (54) |  |  |  |
| Birth weight<sup>b</sup> (g), median (25<sup>th</sup>, 75<sup>th</sup>) |  | 2850 (2600, 3120) |  |  |  |
| Sex: male infants, n (%) |  | 539 (48) |  |  |  |
| Singletons, n (%) |  | 1115 (98) |  |  |  |
| Postnatal hospital stay (days), median (25<sup>th</sup>, 75<sup>th</sup>) |  | 4 (1, 4) |  |  |  |
| Vaginal delivery |  | 1 (1, 1) |  |  |  |
| C-section delivery |  | 4 (4, 5) |  |  |  |
| Infant antibiotic exposure<sup>c</sup>, n (%) |  |  |  |  |  |
| Up to 1 month of age |  | 225 (20) |  |  |  |
| Up to 2 months of age |  | 455 (41) |  |  |  |
| Up to 6 months of age |  | 629 (62) |  |  |  |
| Human milk feeding pattern<sup>d</sup>, n (%) |  | Exclusive | Predominant | Partial | None |
| At or near enrollment |  | 988 (88) | 111 (9.8) | 5 (0.4) | 2 (0.2) |
| At 1 month of age |  | 678 (65) | 217 (21) | 90 (8.0) | 11 (1.1) |
| At 2 months of age |  | 576 (52) | 266 (24) | 28 (2.5) | 22 (2.0) |
| At 6 months of age |  | 79 (8.8) | 70 (7.8) | 232 (20) | 37 (4.1) |
| Diarrheal illness<sup>e</sup>, n (%) |  |  |  |  |  |
| Up to 1 month of age |  | 43 (3.8) |  |  |  |
| Up to 2 months of age |  | 88 (7.9) |  |  |  |
| Up to 6 months of age |  | 195 (19) |  |  |  |
| ***Mother and household characteristics*** |  | ***n = 1125*** |  |  |  |
| Maternal age (years), median (25<sup>th</sup>, 75<sup>th</sup>) |  | 24 (20, 28) |  |  |  |
| Parity, median (25<sup>th</sup>, 75<sup>th</sup>) |  | 2 (1, 2) |  |  |  |
| Primiparous, n (%) |  | 454 (40) |  |  |  |
| Maternal peripartum antibiotics<sup>f</sup>, n (%) |  |  |  |  |  |
| None |  | 57 (5.1) |  |  |  |
| Intrapartum only |  | 65 (5.8) |  |  |  |
| Postpartum only |  | 804 (71) |  |  |  |
| Both intrapartum and postpartum |  | 199 (18) |  |  |  |
| Maternal education, n (%) |  |  |  |  |  |
| Little to no schooling<sup>g</sup> |  | 318 (28) |  |  |  |
| Secondary incomplete |  | 379 (34) |  |  |  |
| Secondary complete or higher |  | 428 (38) |  |  |  |
| Asset index quintile<sup>h</sup>, n (%) |  |  |  |  |  |
| 1 (lowest wealth) |  | 206 (18) |  |  |  |
| 2 |  | 224 (20) |  |  |  |
| 3 |  | 227 (20) |  |  |  |
| 4 |  | 243 (22) |  |  |  |
| 5 (highest wealth) |  | 215 (19) |  |  |  |
| Household size<sup>i</sup>, median (25<sup>th</sup>, 75<sup>th</sup>) |  | 5 (4, 7) |  |  |  |
| One or more household members ≤ 5 years old<sup>j</sup>, n (%) |  | 353 (33) |  |  |  |

<sup>a</sup> Missing data: gestational age (n=6, 0.5%); infant antibiotic exposure (n=13 (1.1%), n=22 (1.9%), n=117 (10%) for 1, 2 and 6 months of age, respectively); human milk feeding pattern (n=5 (0.4%), n=90 (8%), n=28 (2.5%), n=232 (20%), for 1, 2 and 6 months of age, respectively); diarrheal illness (n=13 (1.1%), n=23 (2%), n=116 (10%) for 1, 2 and 6 months of age, respectively); asset index (n=10, 0.9%); household size (n=17, 1.5%); members ≤ 5 years old (n=58, 5.2%).
<sup>b</sup> Weight measured by study personnel was used as a birthweight proxy for four infants where there was >15% difference between birthweight and weight measured within 1-4 days of birth; for three infants, weight was measured within 1 day of birth and, for one infant, weight measured within 4 days of birth was used.
<sup>c</sup> Antibiotic exposure status was derived as any exposure versus no exposure within age windows around the scheduled visits at 1 month (25-31 days), 2 months (46-74 days) and 6 months (173-225 days). Data were considered missing for an infant if their last study visit happened before the age window cut-off.
<sup>d</sup> Human milk feeding pattern was derived based on the infant's feeding status at the closest scheduled visit available at enrollment (0-4 days), and at 1, 2 and 6 months (same age windows as defined in 'c'). Data were considered missing for an infant if there were no study visits within the age window.
<sup>e</sup> Diarrheal illness status was defined as any previous episode of diarrhea (versus never), reported at scheduled visits around 1, 2 and 6 months of age (same age windows and criteria for missing data as defined in 'c').
<sup>f</sup> Intrapartum refers to any antibiotics administered before labour (at hospital admission in which delivery occurred), during labour and/or in the operating theatre, but prior to delivery. Postpartum refers to antibiotics administered after the mother has delivered the infant.
<sup>g</sup> Includes women with no formal education, and incomplete and completed primary school.
<sup>h</sup> Asset index scores were generated using Principal Components Analysis (PCA) for all participants enrolled in the observational cohort study (n=1886) and in a concurrently running trial (n=519) at the same study sites with the same eligibility criteria. The score is a summary measure of household wealth based on ownership of the following assets: electricity, radio, television, almirah, fan, table, chair, fridge, pump, freezer, phone, animals, mobile, watch, computer, autobike, bicycle, rickshaw and a vehicle<sup>65</sup>.
<sup>i</sup> Number of people that sleep in the same house as the infant enrolled in the study, including the infant's twin, if applicable, but excluding the infant enrolled in the study.
<sup>j</sup> Excludes the infant enrolled in the study.

### Abundance of B. infantis, B. longum, and B. breve in the infant stool microbiota

A total of 4613 infant stool samples were tested for *B. infantis* (**Figure 1**). A subset of the samples was tested for *B. longum* (n=1123) and *B. breve* (n=1161) (**Figure 1, Table S3**); 1120 samples (from 364 infants) were tested for all three bacterial targets.

The prevalence of *B. infantis* (i.e., proportion of samples tested in which the bacterial target was detected) and median AA of *B. infantis* (cells/µg DNA) varied with age (**Figure 2A**, **Figure 2B**). Approximately 35% of samples had detectable *B. infantis* during the first week of life (**Figure 2A**), when the median AA was below 10^3^ cells/µg DNA, but AA increased thereafter (**Figure 2B**), such that by 2 months of age, over 60% of the infant stool samples had detectable *B. infantis* (**Figure 2A**) and the median AA was ∼10^8^ cells/µg DNA (**Figure 2B**). High AA persisted to six months of age, when 94% of samples had detectable *B. infantis* (**Figure 2A**, **Figure 2B**).

**Figure 2.**
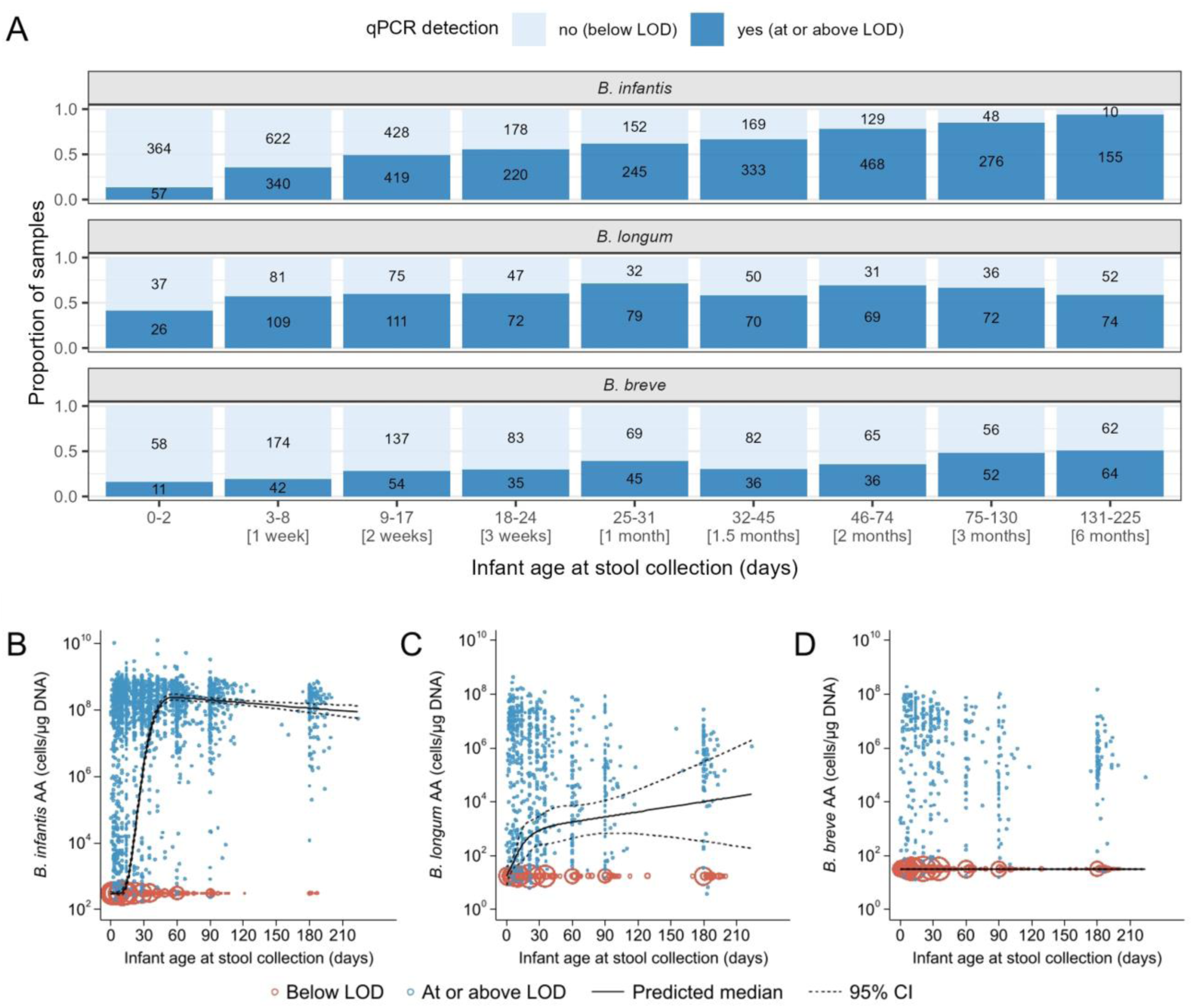
*Bifidobacterium* (sub)species trajectory in early infancy. **(A)** Prevalence and **(B-D)** Absolute abundance of *B. infantis* (n=4613), *B. longum* (n=1123), *B. breve* (n=1161). Sample size (n) refers to the number of samples used in the analysis. The predicted median of absolute abundance (AA) and 95% confidence interval (95% CI) are based on a quantile regression model with clustered standard errors and restricted cubic splines (knots at 7, 14, 28 and 60 days of age). Samples below the assay limit of detection (LOD) were imputed as the median of one-half the LOD normalized to µg DNA. The size of each circle is proportional to the number of overlapping points.

*B. longum* prevalence also varied with age (**Figure 2A**, **Figure 2C**); the median AA increased gradually and was highest (∼10^4^ cells/µg DNA) at the end of the follow-up period (**Figure 2C**). Among the three bacterial targets, *B. breve* had the lowest prevalence overall, with only 16% of samples at 0-2 days of age and 51% of samples by six months of age containing detectable levels (**Figure 2A**); therefore, the median AA of *B. breve* throughout the six-month period was the same as the imputed value that was assigned to all samples below the assay limit of detection (LOD) (**Figure 2D**). The AA of all three bacterial targets exhibited some degree of bimodality from birth to end of the six-month follow-up period (**Figure 3**). While *B. infantis* and *B. breve* had nearly distinct groups of high versus low AAs, this distinction was less clear for *B. longum*.

**Figure 3.**
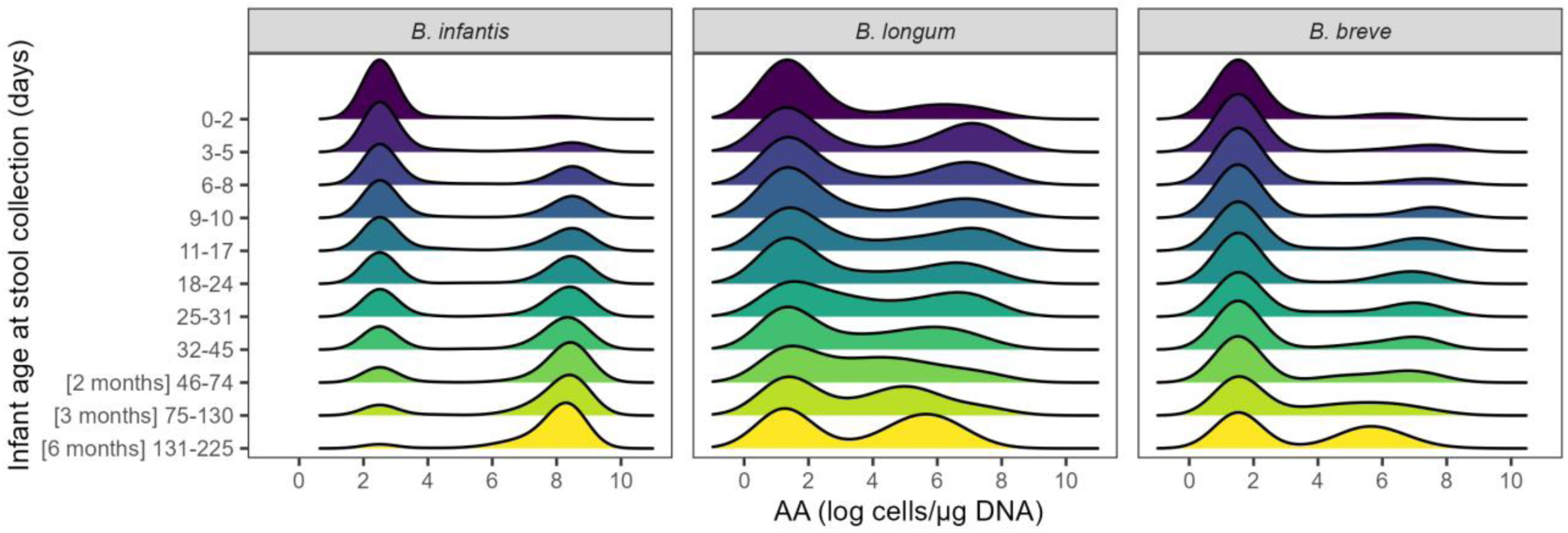
Bimodal distribution of *Bifidobacterium* (sub)species absolute abundance (AA) in infant stool samples, by age windows. *B. infantis* (n=4613); *B. longum* (n=1123); *B. breve* (n=1161). Samples below the assay limit of detection were imputed as the median of one-half the LOD normalized to µg DNA (see Methods for details).

In a sensitivity analysis using different approaches to derive AA (i.e., cell counts normalized to stool mass or not normalized), the AA trajectory within the first six months of age was similar across each of the three methods; differences in magnitude of the AAs were inherent to the scale of each derivation method (**Figure S3, Table S4**).

### B. infantis abundance is inversely related to B. longum and B. breve abundances early in life

We compared the AA, dichotomized as high or low, between *B. infantis* and *B. longum / B. breve*, across different age windows. From birth up to 2.5 months of age, samples with low levels of *B. longum* were more likely to have high abundance of *B. infantis* than samples with high *B. longum* abundance (**Figure 4A**). A similar pattern was observed between *B. infantis* and *B. breve*, with a significant inverse association across all age windows (**Figure 4B**).

**Figure 4.**
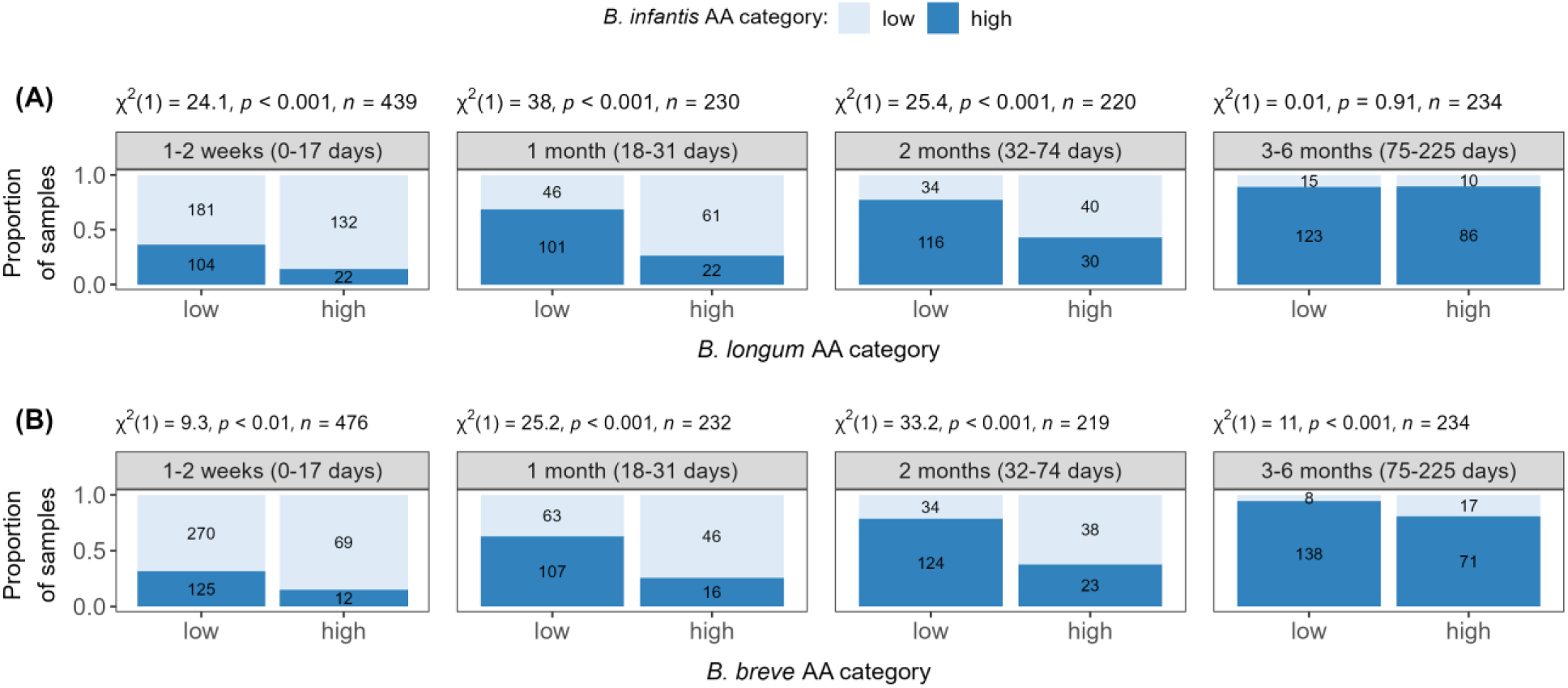
Association of absolute abundance levels between *Bifidobacterium* (sub)species, by age windows. **(A)** *B. infantis* versus *B. longum* (n=1123 samples); **(B)** *B. infantis* versus *B. breve* (n=1161 samples). Categories correspond to low and high absolute abundance (AA), where high AA: ≥6, ≥4.7, ≥4.6 log cells per µg DNA of *B. infantis, B. longum* and *B. breve*, respectively. Number of samples with low/high AA is annotated inside the bars. Pearson chi-square (χ^2^) test results are presented above each individual plot.

### Early emergence of B. infantis is associated with mode of delivery but not infant antibiotic exposure or feeding pattern

The early *B. infantis* AA trajectory differed by mode of delivery (**Figure 5A**). The emergence of a high *B. infantis* AA occurred earlier in infants born by C-section compared to vaginally delivered infants, in which the largest difference observed was at 2-3 weeks of age (∼log 4.5 difference in the AA predicted median) (**Table S5**). Nevertheless, both groups attained a high *B. infantis* abundance by two months of age that persisted to six months of age. This association was not confounded by any maternal characteristics assessed (gestational age, parity, maternal age, maternal education and asset index) (**Figure S4**).

**Figure 5.**
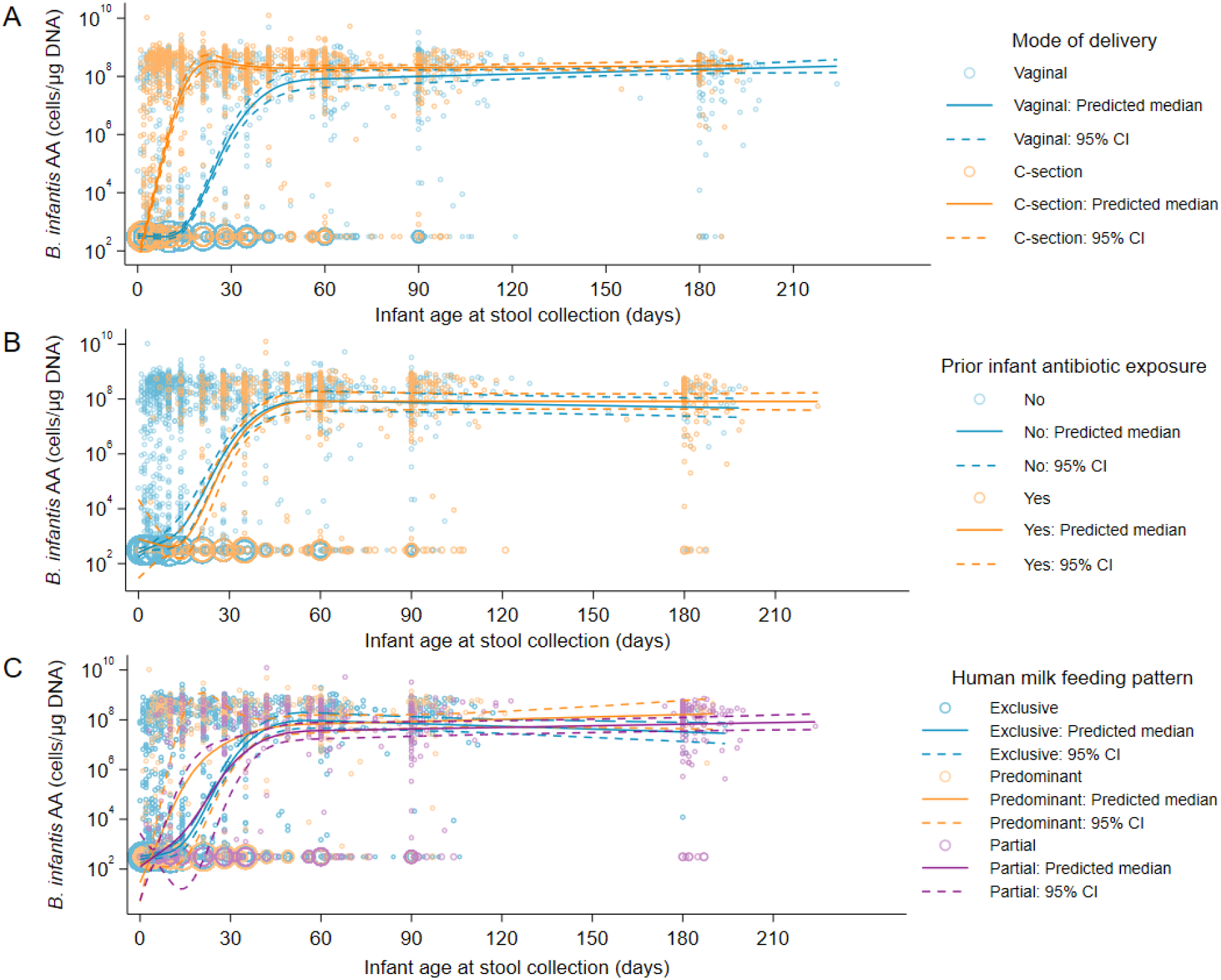
*B. infantis* absolute abundance trajectory, by infant-maternal characteristics. **(A)** Mode of delivery (n=4558); **(B)** Infant antibiotic exposure (n=4558); **(C)** Infant feeding pattern (n=4485). Sample size (n) refers to the number of samples. The predicted median of absolute abundance (AA) and 95% confidence interval (95% CI) are based on a quantile regression model with clustered standard errors and restricted cubic splines (knots at 7, 14, 28 and 60 days of age), adjusted for potential confounders. Samples below the assay limit of detection (LOD) were imputed as the median of one-half the LOD normalized to µg DNA. The size of each circle is proportional to the number of overlapping points. The ‘none’ category for feeding pattern was dropped due to small sample size (n=36 of 4521).

There was a statistically significant positive association of longer postnatal hospital stay with C-section delivery and with higher *B. infantis* AA; therefore, we hypothesized that length of hospital stay could explain the differences in *B. infantis* AA between vaginal and C-section delivery groups within the first month of age. However, the association of mode of delivery with *B. infantis* AA persisted after controlling for the duration of postnatal hospital stay, which was therefore not a mediator ( **Table S6**). The mode of delivery coefficient also did not attenuate after adjusting for maternal exposure to intrapartum antibiotics. Thus, despite being associated with C-section delivery and, to a smaller extent, with *B. infantis* AA, maternal exposure to intrapartum antibiotics was not considered to be a mediator of the association between mode of delivery and *B. infantis* AA (**Table S6**).

Overall, 62% of infants had at least one reported antibiotic exposure during the first six months of age (**Table 1, Figure S5A**); the median infant age at first antibiotic exposure was 46 days. Despite high rates of antibiotic use in infants in this cohort, exposure to antibiotics was not associated with *B. infantis* emergence or AA trajectories within the first six months postnatal age (**Figure 5B**). Overall, the most common systemically administered antibiotics were cefpodoxime, cefixime (both 3^rd^-generation cephalosporins), cefaclor (2^nd^-generation cephalosporin), and amoxicillin, which comprised 51% of all antibiotics reported. At six months of age, cefixime and azithromycin (macrolide) were the most common antibiotics reported (**Figure S5B**).

Overall, most infants received some human milk throughout their first six months after delivery (**Table 1, Figure S5C-E**), yet differences in feeding patterns, irrespective of whether other liquids/foods were consumed in addition to or besides human milk, were not associated with the trajectory of *B. infantis* AA (**Figure 5C**). Among infants who were not receiving human milk exclusively, the most common food consumed at 1-3 months of age was infant formula, followed by honey/date-based syrup and plain water. At six months of age, the most common foods reported were complementary solid foods, plain water and infant formula (**Figure S5D**).

Infant antibiotic exposure and feeding pattern were also identified as candidate mediators of the association between mode of delivery and *B. infantis* AA, but since these variables were not associated with *B. infantis* AA, mediation analyses were not performed as they would be unlikely to explain differences in AA by mode of delivery.

A sensitivity analysis restricted to participants enrolled in schedules A and B only (see Data and sample collection in Methods for more details) was conducted due to the possible effects of the COVID-19 pandemic, calendar date, and different longitudinal stool sample sizes on *B. infantis* AA; inferences were similar to the primary analysis for all three exposures of interest ( **Figure S6**). The associations of each exposure of interest with the probabilities of samples having high *B. infantis* AA were also estimated (**Figure S7**). Stool samples obtained from infants born by C-section had a higher probability of high *B. infantis* AA compared to vaginally delivered infants, particularly in the first two months of age. Consistent with the primary analysis, infant antibiotic exposure and feeding pattern were not associated with the probability of high AA at any age (**Figure S7**).

### Relative abundance trajectory of B. longum subspecies (longum and infantis) corroborates that B. infantis is highly abundant and dominant in this population

*B. infantis* relative abundance (RA) increased sharply within the first month of age among infants born by C-section, remaining high (∼80%) throughout the whole six-month follow-up period (**Figure 6**), similar to the pattern observed for *B. infantis* AA. Among vaginally delivered infants, *B. infants* RA also increased with age with a peak of 45% by two months of age that was maintained until the end of the six-month follow-up period. *B. longum* RA was low for both modes of delivery (**Figure 6**), which was similar to the corresponding AA trajectory.

**Figure 6.**
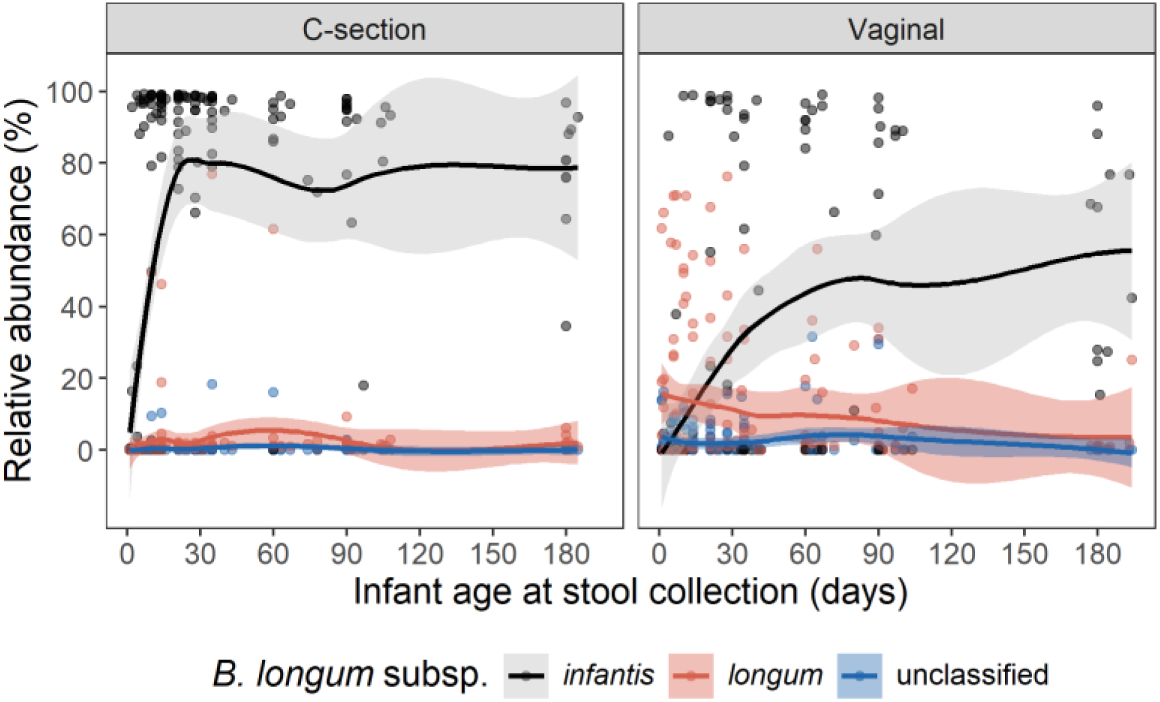
*B. longum* subspecies (*longum* and *infantis*) relative abundance trajectory, by mode of delivery. Relative abundance (RA) is shown as a loess curve (solid line) and 95% confidence interval (shade) based on metagenomic analysis using MetaPhlAn4, restricted to samples from schedule A (n=55 infants, n=332 samples). Values refer to the RA of each target shown in relation to the complete microbiota profile (i.e., all bacterial taxa).

### Maternal stool B. infantis may not contribute to seeding the newborn gut microbiota

We investigated whether the maternal stool microbiota during the first week postpartum could be a potential source of *B. infantis* for seeding the newborn gut microbiota. Approximately 25% of mothers (126/491) had *B. infantis* detected in their stool samples at 0-7 days postpartum, which was associated with infant *B. infantis* AA in the unadjusted model, in the first month after delivery (**Table S7**). However, after controlling for mode of delivery, maternal *B. infantis* detection was no longer associated with infant *B. infantis* AA. We further explored whether this was confounded by underlying factors potentially associated with both C-section delivery and maternal *B. infantis* detection (e.g., socioeconomic factors); however, neither asset index (p=0.37) nor maternal education (p=0.12) was associated with the detection of B. infantis in maternal stool samples.

## DISCUSSION

Bifidobacteria are generally considered typical members of a healthy gut microbiota in early infancy^2^, but several factors may influence the timing of their emergence, overall abundance and (sub)species dominance patterns. In an urban cohort of infants in Bangladesh, *B. infantis*, *B. longum,* and *B. breve* had distinct abundance trajectories during the first six months after delivery. *B. infantis* was not frequently detected at birth but became widespread and reached high abundance at an early age (∼1 month), assessed by qPCR and corroborated by metagenomics. Considering that *B. longum* and *B. breve* had low abundance overall in this population, we focused on *B. infantis* to investigate various infant-maternal characteristics that could be associated with its emergence and abundance trajectory.

Consistent with other studies, infants from LMICs typically have a gut microbiota dominated by bifidobacteria, and particularly *B. infantis* (Bangladesh^23,24,25^; Gambia^26^; Indonesia^27^; Pakistan and Tanzania^28^). Conversely, some populations from high income countries (HIC) have either low abundance or no *B. infantis* (Belgium^29^; Canada^30^; Japan^31^; New Zealand^27^; Singapore^32^; USA^33,34^; UK^35^). The *B. infantis* AA peak at 1-2 months is similar to the pattern described by Barrat et al.^24^ for healthy infants residing in an urban community in Dhaka.

The AA distributions of *B. infantis, B. longum* and *B. breve* were bimodal indicating that the microbiota composition undergoes rapid changes, and that an intermediate abundance level may not be frequently observed, despite longitudinal sampling. A similar distribution has been reported by others for *Bifidobacterium*^36,37^ and *B. infantis*^28^, irrespective of the quantification method used (absolute or relative abundance). Bimodality has also been observed in other gut taxa of an adult population, where bacteria were either highly abundant or nearly absent^38^. This phenomenon has been linked to the alternative stable states theory, which suggests that stable states (e.g., low and high abundance) are separated by an unstable tipping point (intermediate abundance) that may lead to abrupt shifts in the microbial composition upon even small fluctuations in the ecosystem (e.g., induced by microbial competition, host environmental factors and others)^38^.

To better understand the ecological dynamics involving the early emergence of a high abundance state of *B. infantis*, its abundance was compared against the other two targets, which showed a negative association. Although causality cannot be inferred from this analysis, the results suggest competition between *B. infantis* and *B. longum,* and *B. infantis* and *B. breve,* corroborating findings from other studies^28,17^. Based on the longitudinal pattern of prevalence and abundance in the first days after delivery, we hypothesize that upon postnatal entry of *B. infantis* in the ecosystem (since it was rare at birth), it progressively displaced other *Bifidobacterium* (sub)species.

It is not surprising that *B. infantis* would have a fitness advantage over other bacterial taxa since most infants in this study were exclusively breastfed up to two months of age and *B. infantis* is known for its capacity to utilize HMOs^39^. Nevertheless, *B. infantis* is not always a dominant taxon in populations where human milk feeding is common. The current initiation and exclusive breastfeeding rates in Canada and the US at two months of age, for example, are relatively similar to this study population^40,41^, yet, studies from these countries have shown an overall low abundance of *B. infantis*^30,33^. Taft et al.^42^ proposed that *B. infantis* may be lost from populations with a history of lower breastfeeding rates, irrespective of current rates. Moreover, exclusive human milk feeding has many short- and long-term benefits^43^ and is recommended for the first six months of life^44^, but our results indicate that even low levels of human milk exposure might be sufficient to support *B. infantis* colonization, provided that other people in the local community serve as a reservoir of *B. infantis* and have opportunities to transmit it to newborns.

Mode of delivery was a key factor associated with *B. infantis* abundance, demonstrated by a delayed emergence of *B. infantis* in vaginally delivered babies. These results were consistent with a previous study from Bangladesh in which *Bifidobacterium* and *B. infantis* relative abundances were lower in infants delivered vaginally at six weeks of age in comparison with those delivered by C-section^45^. Vaginal delivery has been generally associated with higher prevalence/abundance of *Bifidobacterium* in comparison to C-section, particularly within the first months of age^29,30,31,32,46,47^. In addition, only a few studies have examined this association with *B. infantis* specifically, perhaps due to its overall low abundance in some infant populations; and, while some reports showed a higher prevalence/abundance of *B. infantis* in vaginally delivered infants (similar pattern often reported for *Bifidobacterium*)^31,48^, others found no association with mode of delivery^30,32^. Importantly, most of these studies are from HIC, and their findings may not be generalized to all populations worldwide. Moreover, while it is possible that C-section may have short- and long-term effects on other bacterial taxa and/or clinical health implications^49^, we found that any differences in *B. infantis* absolute abundance between vaginal and C-section groups attenuated after one month of age and disappeared by six months of age.

We hypothesized that the delay in *B. infantis* emergence in vaginally delivered infants may be explained by the additional microbial exposure from the birth canal typical of a vaginal birth but not C-section. Thus, the lack of some microbes among infants born by C-section may create an environment where *B. infantis* thrives, whereas in infants born by vaginal delivery, there is greater competition between *B. infantis* and other bacterial taxa in the early postnatal period. Using metagenomics in a subset of samples, we confirmed that *B. infantis* was not only abundant but was also a dominant taxon in this population. The RA of *B. infantis* differed by delivery mode (higher RA in the C-section group vs. vaginal) whereas the AA was similar in both groups after two months of age; this suggests that vaginally delivered infants had a higher total bacterial load than infants born by C-section, consistent with previous studies^29,50^. This may relate to the additional microbial exposure during vaginal birth compared to C-section birth, as mentioned above, such that *B. infantis* was able to reach its growth limit but other taxa were also able to successfully colonize in the intestinal tract of the infant.

The maternal microbiota plays an important role in seeding the newborn gut, with delivery mode as one of the main modulators of this event^51,52^, yet the lack of an association between maternal *B. infantis* detection and newborn *B. infantis* abundance suggested that other individuals contributed to *B. infantis* seeding (e.g., siblings). Given that horizontal transmission may play an important role in maintaining *B. infantis* in this population, fewer in-person social interactions as a result of nationwide COVID-19 pandemic public health measures that took place in Bangladesh during the implementation of this study could have impacted *B. infantis* abundance. Our analyses restricted to infants recruited over a shorter interval (i.e., schedules A and B: May-Oct 2021 vs. schedule C: Nov 2020-Oct 2021) showed the same age-related patterns of *B. infantis* emergence and abundance, even though some enrollment occurred during a period in which there were stringent nationwide public health measures (i.e., various intervals from June 26 to August 5, 2021).

Except for amoxicillin, the antibiotics most commonly administered to infants in this study (i.e., 2^nd^ or 3^rd^ generation cephalosporines, and azithromycin) are in the ‘watch’ category of the WHO AWaRe (Access, Watch, Reserve) classification^53^. Antibiotic resistance is recognized as a major public health issue^54^, and ‘watch’ antibiotics have higher resistance potential, so it has been recommended that their use be monitored to ensure appropriate stewardship^55^. There was no association between *B. infantis* abundance and infant antibiotic exposure, but it is unlikely that the prevalent *B. infantis* strains were resistant to these drugs and it may suggest they were able to rapidly recolonize the infant intestinal tract upon antibiotic discontinuation. Antimicrobials often associated with resistance in bifidobacteria (e.g., mupirocin, tetracycline) ^56,57^ were not among the top systemic antibiotics reported. Bifidobacteria are generally susceptible to cephalosporins^58^, amoxicillin^58,59^, and ampicillin^60,61^, which were commonly reported in this study, particularly within the first month of age; they are also generally susceptible to macrolides like azithromycin, which was mainly reported at six months of age^62,63^. Bifidobacteria resistance varies against aminoglycosides, like amikacin and gentamycin^58,60,61^, which were reported mainly during the first month of age but represented a small proportion of the total number of antibiotics administered at this time point.

A limitation of this study was the lower representation of some clinical populations (e.g., low birth weight, preterm infants) due to the eligibility criteria applied (described in the Methods), which were intended to be applicable to a postnatal intervention trial. Also, the small number of infants who were not exposed to any human milk underpowered our ability to investigate the potential association between no breastfeeding and *B. infantis* abundance. Moreover, considering that maternal samples were not collected prior to birth, we could not determine with certainty whether mode of delivery had a direct effect on the maternal stool *B. infantis*, or other unknown underlying factors were implicated in this association. Lastly, the antimicrobial susceptibility patterns reported in the literature could not be empirically confirmed since bacteria isolates were not cultured from stool samples (for use in testing of antimicrobial susceptibility) nor were any molecular methods used to identify genes associated with resistance; moreover, it has been proposed that breastfeeding may mitigate the effects of antibiotics on the intestinal microbiota^64^, but this could not be investigated due to the low numbers of infants who were not breastfed.

In conclusion, there are distinct patterns of emergence of typical members of the infant gut microbiota (*B. infantis, B. longum* and *B. breve*) in the early postnatal period of an urban South Asian LMIC infant population. By differentiating the major *B. longum* subspecies (*longum* and *infantis)*, we were able to describe patterns that have been masked in previous studies based on 16S rRNA sequencing or metagenomic profiling. The large sample size and longitudinal study design enable precise tracking of the abundance trajectories and estimation of associations of *B. infantis* abundance with selected infant-maternal characteristics. Contrary to observations based on HIC study populations, we observed a delay in intestinal colonization of *B. infantis* in vaginally delivered infants. Nevertheless, by two months of age, *B. infantis* achieved high abundance levels in nearly all infant samples, irrespective of their mode of delivery. Being exposed to antibiotics or having a mixed breastfeeding pattern did not have an impact on *B. infantis* abundance in this population. Lastly, probiotics including several *B. infantis* strains have been a focus of substantial interest as a potential strategy to reduce the burden of bacterial infections and neonatal mortality rates in South Asian countries. However, the high prevalence and abundance of *B. infantis* highlights the need for careful selection of appropriate probiotic species/strains best suited to various target populations (e.g., healthy vs. preterm, infants with severe acute malnutrition, etc.). Given that clinical benefits either from commensals or probiotics are likely strain-specific, future work will aim to identify specific *B. infantis* strains and their genomic features that account for the postnatal colonization observed in this population.

## Supporting information

Freitas_et_al_Bifidobacterium_Supplemental_material.zip

## ACKNOWLEDGMENTS

We thank the families who were involved in the study and all SEPSiS co-investigators and study personnel who contributed to the SEPSiS observational cohort study. We thank Samir Kumar Saha (Child Health Research Foundation, Bangladesh) for supervision of field operations, Rob Knight and lab members (UCSD, US) Caitlin Tribelhorn, Sawyer Farmer, Lauren Hansen, Helena Tubb, and Madison Ambre for generating the raw metagenomics data and host-filtering.

## DATA AVAILABILITY

All study materials have been deposited at Borealis and are publicly available as of the date of publication: data and code used in analysis (doi.org/10.5683/SP3/UDMXA2); Standard Operating Procedures (SOPs) used during sample and data collection (doi.org/10.5683/SP3/WKDQYY); Statistical Analysis Plan (doi.org/10.5683/SP3/JEDIJY).

## AUTHOR CONTRIBUTIONS

Conceptualization: L.G.P.; D.G.B.; A.A.M.; D.H.H.; E.P.; M.I.H.; M.M.S.; M.S.I.; P.M.S.; P.S.S.; S.S.; S.K.M.; T.A.; R.H.; S.A.S.; D.E.R. Formal analysis and Visualization: A.C.F.; G.L.; H.Q.; C.H. Investigation: A.C.F.; G.L.; J.S. Project administration: A.C.F.; O.H.O.; S.P-B.; J.C.O.; L.G.P.; K.M.O.C.; D.E.R. Data curation: A.C.F.; G.L. Field or lab activities supervision: M.K.; A.A.M.; M.I.H.; M.S.I.; S.M.A.G.; S.S. Project supervision: R.H.; S.A.S. (the International Centre for Diarrhoeal Disease Research, Bangladesh site); L.G.P.; K.M.O.C.; D.E.R. (the Hospital for Sick Children site). Writing – original draft: A.C.F. Writing – review & editing: G.L.; H.Q.; L.G.P.; O.H.O.; S.P-B.; D.G.B.; K.M.O.C.; J.C.O.; M.G.L.; C.H.; C.W.E.S; M.S.I.; P.M.S.; P.S.S.; S.M.A.G.; S.K.M.; R.H.; D.E.R. Funding acquisition: D.E.R.

## SUPPLEMENTAL MATERIAL

Freitas_et_al_Bifidobacterium_Supplemental_material.pdf (Figures S1–S7 and Tables S1–S7).

## REFERENCES

1. Underwood MA, Mukhopadhyay S, Lakshminrusimha S, Bevins CL. 2020. Neonatal intestinal dysbiosis. J Perinatol. 40(11):1597–1608. doi:10.1038/s41372-020-00829-2

2. Milani C, Duranti S, Bottacini F, Casey E, Turroni F, Mahony J, Belzer C, Delgado Palacio S, Arboleya Montes S, Mancabelli L, et al. 2017. The first microbial colonizers of the human gut: composition, activities, and health implications of the infant gut microbiota. Microbiol Mol Biol Rev. 81(4):e00036–17. doi:10.1128/MMBR.00036-17

3. Bergmann KR, Liu SXL, Tian R, Kushnir A, Turner JR, Li H-L, Chou PM, Weber CR, De Plaen IG. 2013. *Bifidobacteria* stabilize claudins at tight junctions and prevent intestinal barrier dysfunction in mouse necrotizing enterocolitis. The American Journal of Pathology. 182(5):1595–1606. doi:10.1016/j.ajpath.2013.01.013

4. Tsukuda N, Yahagi K, Hara T, Watanabe Y, Matsumoto H, Mori H, Higashi K, Tsuji H, Matsumoto S, Kurokawa K, Matsuki T. 2021. Key bacterial taxa and metabolic pathways affecting gut short -chain fatty acid profiles in early life. ISME J. 15(9):2574–2590. doi:10.1038/s41396-021-00937-7

5. Huda MN, Lewis Z, Kalanetra KM, Rashid M, Ahmad SM, Raqib R, Qadri F, Underwood MA, Mills DA, Stephensen CB. 2014. Stool microbiota and vaccine responses of infants. Pediatrics. 134(2):e362– e372. doi:10.1542/peds.2013-3937

6. Underwood MA, German JB, Lebrilla CB, Mills DA. 2015. *Bifidobacterium longum* subspecies *infantis*: champion colonizer of the infant gut. Pediatr Res. 77(1):229–235. doi:10.1038/pr.2014.156

7. Abdill RJ, Adamowicz EM, Blekhman R. 2022. Public human microbiome data are dominated by highly developed countries. PLOS Biology. 20(2):e3001536. doi:10.1371/journal.pbio.3001536

8. Haarman M, Knol J. 2005. Quantitative real-time PCR assays to identify and quantify fecal *Bifidobacterium* species in infants receiving a prebiotic infant formula. Applied and Environmental Microbiology. 71(5):2318–2324. doi:10.1128/AEM.71.5.2318-2324.2005

9. Lawley B, Munro K, Hughes A, Hodgkinson AJ, Prosser CG, Lowry D, Zhou SJ, Makrides M, Gibson RA, Lay C, et al. 2017. Differentiation of *Bifidobacterium longum* subspecies *longum* and *infantis* by quantitative PCR using functional gene targets. PeerJ. 5:e3375. doi:10.7717/peerj.3375

10. Brennan C, Salido RA, Belda-Ferre P, Bryant M, Cowart C, Tiu MD, González A, McDonald D, Tribelhorn C, Zarrinpar A, Knight R. 2023. Maximizing the potential of high-throughput next-generation sequencing through precise normalization based on read count distribution. mSystems. 8(4):e00006–23. doi:10.1128/msystems.00006-23

11. Gonzalez A, Navas-Molina JA, Kosciolek T, McDonald D, Vázquez-Baeza Y, Ackermann G, DeReus J, Janssen S, Swafford AD, Orchanian SB, et al. 2018. Qiita: rapid, web-enabled microbiome meta-analysis. Nat Methods. 15(10):796–798. doi:10.1038/s41592-018-0141-9

12. Chen S, Zhou Y, Chen Y, Gu J. 2018. fastp: an ultra-fast all-in-one FASTQ preprocessor. Bioinformatics. 34(17):i884–i890. doi:10.1093/bioinformatics/bty560

13. Li H. 2018. Minimap2: pairwise alignment for nucleotide sequences.Birol I, editor. Bioinformatics. 34(18):3094–3100. doi:10.1093/bioinformatics/bty191

14. Liao W-W, Asri M, Ebler J, Doerr D, Haukness M, Hickey G, Lu S, Lucas JK, Monlong J, Abel HJ, et al. 2023. A draft human pangenome reference. Nature. 617(7960):312–324. doi:10.1038/s41586-023-05896-x

15. Zakeri M, Brown NK, Ahmed OY, Gagie T, Langmead B. 2024. Movi: A fast and cache-efficient full-text pangenome index. iScience. 27(12):111464. doi:10.1016/j.isci.2024.111464

16. Blanco-Míguez A, Beghini F, Cumbo F, McIver LJ, Thompson KN, Zolfo M, Manghi P, Dubois L, Huang KD, Thomas AM, et al. 2023. Extending and improving metagenomic taxonomic profiling with uncharacterized species using MetaPhlAn 4. Nat Biotechnol. 41(11):1633–1644. doi:10.1038/s41587-023-01688-w

17. Ennis D, Shmorak S, Jantscher-Krenn E, Yassour M. 2024. Longitudinal quantification of *Bifidobacterium longum* subsp. *infantis* reveals late colonization in the infant gut independent of maternal milk HMO composition. Nat Commun. 15(1):894. doi:10.1038/s41467-024-45209-y

18. Machado J a. F, Parente PMDC, Silva JMCS. 2021. QREG2: Stata module to perform quantile regression with robust and clustered standard errors. Statistical Software Components. S457369.

19. Baron RM, Kenny DA. 1986. The moderator-mediator variable distinction in social psychological research: conceptual, strategic, and statistical considerations. J Pers Soc Psychol. 51(6):1173–1182. doi:10.1037//0022-3514.51.6.1173

20. Patil I. 2021. Visualizations with statistical details: The “ggstatsplot” approach. JOSS. 6(61):3167. doi:10.21105/joss.03167

21. Højsgaard S, Halekoh U, Yan J, Ekstrøm CT. 2024. geepack: Generalized Estimating Equation package. https://cran.r-project.org/web/packages/geepack/index.html

22. Arel-Bundock V, Greifer N, Bacher E, McDermott G, Heiss A. 2024. marginaleffects: Predictions, comparisons, slopes, marginal means, and hypothesis tests. https://cran.r-project.org/web/packages/marginaleffects/index.html

23. Huda MN, Ahmad SM, Alam MJ, Khanam A, Kalanetra KM, Taft DH, Raqib R, Underwood MA, Mills DA, Stephensen CB. 2019. *Bifidobacterium* abundance in early infancy and vaccine response at 2 years of age. Pediatrics. 143(2):e20181489. doi:10.1542/peds.2018-1489

24. Barratt MJ, Nuzhat S, Ahsan K, Frese SA, Arzamasov AA, Sarker SA, Islam MM, Palit P, Islam MR, Hibberd MC, et al. 2022. *Bifidobacterium infantis* treatment promotes weight gain in Bangladeshi infants with severe acute malnutrition. Sci Transl Med. 14(640):eabk1107. doi:10.1126/scitranslmed.abk1107

25. Vatanen T, Ang QY, Siegwald L, Sarker SA, Le Roy CI, Duboux S, Delannoy-Bruno O, Ngom-Bru C, Boulangé CL, Stražar M, et al. 2022. A distinct clade of *Bifidobacterium longum* in the gut of Bangladeshi children thrives during weaning. Cell. 185(23):4280–4297.e12. doi:10.1016/j.cell.2022.10.011

26. Davis JCC, Lewis ZT, Krishnan S, Bernstein RM, Moore SE, Prentice AM, Mills DA, Lebrilla CB, Zivkovic AM. 2017. Growth and morbidity of Gambian infants are influenced by maternal milk oligosaccharides and infant gut microbiota. Sci Rep. 7(1):40466. doi:10.1038/srep40466

27. Lawley B, Otal A, Moloney-Geany K, Diana A, Houghton L, Heath A-LM, Taylor RW, Tannock GW. 2019. Fecal microbiotas of Indonesian and New Zealand children differ in complexity and bifidobacterial taxa during the first year of life.Elkins CA, editor. Appl Environ Microbiol. 85(19):e01105–19. doi:10.1128/AEM.01105-19

28. Colston JM, Taniuchi M, Ahmed T, Ferdousi T, Kabir F, Mduma E, Nshama R, Iqbal NT, Haque R, Ahmed T, et al. 2022. Intestinal colonization with *Bifidobacterium longum* subspecies is associated with length at birth, exclusive breastfeeding, and decreased risk of enteric virus infections, but not with histo-blood group antigens, oral vaccine response or later growth in three birth cohorts. Front Pediatr. 10:804798. doi:10.3389/fped.2022.804798

29. Martin R, Makino H, Yavuz AC, Ben-Amor K, Roelofs M, Ishikawa E, Kubota H, Swinkels S, Sakai T, Oishi K, et al. 2016. Early-life events, including mode of delivery and type of feeding, siblings and gender, shape the developing gut microbiota. PLOS ONE. 11(6):e0158498. doi:10.1371/journal.pone.0158498

30. Chen YY, Zhao X, Moeder W, Tun HM, Simons E, Mandhane PJ, Moraes TJ, Turvey SE, Subbarao P, Scott JA, Kozyrskyj AL. 2021. Impact of maternal intrapartum antibiotics, and caesarean section with and without labour on *Bifidobacterium* and other infant gut microbiota. Microorganisms. 9(9):1847. doi:10.3390/microorganisms9091847

31. Nagpal R, Kurakawa T, Tsuji H, Takahashi T, Kawashima K, Nagata S, Nomoto K, Yamashiro Y. 2017. Evolution of gut *Bifidobacterium* population in healthy Japanese infants over the first three years of life: a quantitative assessment. Sci Rep. 7(1):10097. doi:10.1038/s41598-017-10711-5

32. Xu J, Duar RM, Quah B, Gong M, Tin F, Chan P, Sim CK, Tan KH, Chong YS, Gluckman PD, et al. 2024. Delayed colonization of *Bifidobacterium* spp. and low prevalence of B. infantis among infants of Asian ancestry born in Singapore: insights from the GUSTO cohort study. Front Pediatr. 12:1421051. doi:10.3389/fped.2024.1421051

33. Frese SA, Hutton AA, Contreras LN, Shaw CA, Palumbo MC, Casaburi G, Xu G, Davis JCC, Lebrilla CB, Henrick BM, et al. 2017. Persistence of supplemented *Bifidobacterium longum* subsp. *infantis* EVC001 in breastfed infants. mSphere. 2(6):10.1128/msphere.00501-17. doi:10.1128/msphere.00501-17

34. Casaburi G, Duar RM, Brown H, Mitchell RD, Kazi S, Chew S, Cagney O, Flannery RL, Sylvester KG, Frese SA, et al. 2021. Metagenomic insights of the infant microbiome community structure and function across multiple sites in the United States. Sci Rep. 11(1):1472. doi:10.1038/s41598-020-80583-9

35. Shao Y, Garcia-Mauriño C, Clare S, Dawson NJR, Mu A, Adoum A, Harcourt K, Liu J, Browne HP, Stares MD, et al. 2024. Primary succession of Bifidobacteria drives pathogen resistance in neonatal microbiota assembly. Nat Microbiol.:1–13. doi:10.1038/s41564-024-01804-9

36. Lewis ZT, Totten SM, Smilowitz JT, Popovic M, Parker E, Lemay DG, Van Tassell ML, Miller MJ, Jin Y-S, German JB, et al. 2015. Maternal fucosyltransferase 2 status affects the gut bifidobacterial communities of breastfed infants. Microbiome. 3(1):13. doi:10.1186/s40168-015-0071-z

37. Taft DH, Liu J, Maldonado-Gomez MX, Akre S, Huda MN, Ahmad SM, Stephensen CB, Mills DA. 2018. Bifidobacterial dominance of the gut in early life and acquisition of antimicrobial resistance. mSphere. 3(5):e00441–18. doi:10.1128/mSphere.00441-18

38. Lahti L, Salojärvi J, Salonen A, Scheffer M, de Vos WM. 2014. Tipping elements in the human intestinal ecosystem. Nat Commun. 5(1):4344. doi:10.1038/ncomms5344

39. Sela DA, Chapman J, Adeuya A, Kim JH, Chen F, Whitehead TR, Lapidus A, Rokhsar DS, Lebrilla CB, German JB, et al. 2008. The genome sequence of *Bifidobacterium longum* subsp. *infantis* reveals adaptations for milk utilization within the infant microbiome. Proceedings of the National Academy of Sciences. 105(48):18964–18969. doi:10.1073/pnas.0809584105

40. CDC. 2022. (Centers for Disease Control and Prevention) Breastfeeding report card, United States. https://www.cdc.gov/breastfeeding/pdf/2022-breastfeeding-report-card-h.pdf

41. PHAC. 2022. (Public Health Agency of Canada). Canada’s breastfeeding progress report. https://health-infobase.canada.ca/src/data/breastfeeding/PHAC%20-%20Breastfeeding%20Report%202022.pdf

42. Taft DH, Lewis ZT, Nguyen N, Ho S, Masarweh C, Dunne-Castagna V, Tancredi DJ, Huda MN, Stephensen CB, Hinde K, et al. 2022. *Bifidobacterium* species colonization in infancy: A global cross-sectional comparison by population history of breastfeeding. Nutrients. 14(7):1423. doi:10.3390/nu14071423

43. Victora CG, Bahl R, Barros AJD, França GVA, Horton S, Krasevec J, Murch S, Sankar MJ, Walker N, Rollins NC. 2016. Breastfeeding in the 21st century: epidemiology, mechanisms, and lifelong effect. The Lancet. 387(10017):475–490. doi:10.1016/S0140-6736(15)01024-7

44. WHO. 2023. (World Health Organization) Infant and young child feeding. [accessed 2024 Dec 16]. https://www.who.int/news-room/fact-sheets/detail/infant-and-young-child-feeding

45. Huda MN, Ahmad SM, Kalanetra KM, Taft DH, Alam MJ, Khanam A, Raqib R, Underwood MA, Mills DA, Stephensen CB. 2019. Neonatal vitamin A supplementation and vitamin A status are associated with gut microbiome composition in Bangladeshi infants in early infancy and at 2 years of age. The Journal of Nutrition. 149(6):1075. doi:10.1093/jn/nxz034

46. Reyman M, van Houten MA, van Baarle D, Bosch AATM, Man WH, Chu MLJN, Arp K, Watson RL, Sanders EAM, Fuentes S, Bogaert D. 2019. Impact of delivery mode-associated gut microbiota dynamics on health in the first year of life. Nat Commun. 10(1):4997. doi: 10.1038/s41467-019-13014-7

47. Shao Y, Forster SC, Tsaliki E, Vervier K, Strang A, Simpson N, Kumar N, Stares MD, Rodger A, Brocklehurst P, et al. 2019. Stunted microbiota and opportunistic pathogen colonization in caesarean-section birth. Nature. 574(7776):117–121. doi:10.1038/s41586-019-1560-1

48. Seppo AE, Bu K, Jumabaeva M, Thakar J, Choudhury RA, Yonemitsu C, Bode L, Martina CA, Allen M, Tamburini S, et al. 2021. Infant gut microbiome is enriched with *Bifidobacterium longum* ssp. *infantis* in Old Order Mennonites with traditional farming lifestyle. Allergy. 76(11):3489–3503. doi:10.1111/all.14877

49. Sandall J, Tribe RM, Avery L, Mola G, Visser GH, Homer CS, Gibbons D, Kelly NM, Kennedy HP, Kidanto H, et al. 2018. Short-term and long-term effects of caesarean section on the health of women and children. The Lancet. 392(10155):1349–1357. doi:10.1016/S0140-6736(18)31930-5

50. Selma-Royo M, Calatayud Arroyo M, García-Mantrana I, Parra-Llorca A, Escuriet R, Martínez-Costa C, Collado MC. 2020. Perinatal environment shapes microbiota colonization and infant growth: impact on host response and intestinal function. Microbiome. 8( 1):167. doi:10.1186/s40168-020-00940-8

51. Bogaert D, van Beveren GJ, de Koff EM, Lusarreta Parga P, Balcazar Lopez CE, Koppensteiner L, Clerc M, Hasrat R, Arp K, Chu MLJN, et al. 2023. Mother-to-infant microbiota transmission and infant microbiota development across multiple body sites. Cell Host & Microbe. 31(3):447–460.e6. doi:10.1016/j.chom.2023.01.018

52. Selma-Royo M, Dubois L, Manara S, Armanini F, Cabrera-Rubio R, Valles-Colomer M, González S, Parra-Llorca A, Escuriet R, Bode L, et al. 2024. Birthmode and environment-dependent microbiota transmission dynamics are complemented by breastfeeding during the first year. Cell Host & Microbe. 32(6):996–1010.e4. doi:10.1016/j.chom.2024.05.005

53. WHO. 2023. (World Health Organization) AWaRe (access, watch, reserve) classification of antibiotics for evaluation and monitoring of use, 2023. In: The selection and use of essential medicines 2023: Executive summary of the report of the 24^th^ WHO Expert Committee on the Selection and Use of Essential Medicines [Internet]. Geneva: World Health Organization. https://www.who.int/publications/i/item/WHO-MHP-HPS-EML-2023.04

54. Naghavi M, Vollset SE, Ikuta KS, Swetschinski LR, Gray AP, Wool EE, Aguilar GR, Mestrovic T, Smith G, Han C, et al. 2024. Global burden of bacterial antimicrobial resistance 1990–2021: a systematic analysis with forecasts to 2050. The Lancet. 404(10459):1199–1226. doi:10.1016/S0140-6736(24)01867-1

55. Zanichelli V, Sharland M, Cappello B, Moja L, Getahun H, Pessoa-Silva C, Sati H, Weezenbeek C van, Balkhy H, Simão M, et al. 2023. The WHO AWaRe (Access, Watch, Reserve) antibiotic book and prevention of antimicrobial resistance. Bulletin of the World Health Organization. 101(4):290. doi:10.2471/BLT.22.288614

56. Serafini F, Bottacini F, Viappiani A, Baruffini E, Turroni F, Foroni E, Lodi T, Sinderen D van, Ventura M. 2011. Insights into physiological and genetic mupirocin susceptibility in bifidobacteria. Applied and Environmental Microbiology. 77(9):3141. doi:10.1128/AEM.02540-10

57. Gueimonde M, Sánchez B, G. De Los Reyes-Gavilán C, Margolles A. 2013. Antibiotic resistance in probiotic bacteria. Front Microbiol. 4. doi:10.3389/fmicb.2013.00202

58. Yazid A m., Ali A m., Shuhaimi M, Kalaivaani V, Rokiah M y., Reezal A. 2000. Antimicrobial susceptibility of bifidobacteria. Letters in Applied Microbiology. 31(1):57–62. doi:10.1046/j.1472-765x.2000.00764.x

59. Delgado S, Flórez AB, Mayo B. 2005. Antibiotic susceptibility of *Lactobacillus* and *Bifidobacterium* species from the human gastrointestinal tract. Curr Microbiol. 50(4):202–207. doi:10.1007/s00284-004-4431-3

60. Duranti S, Lugli GA, Mancabelli L, Turroni F, Milani C, Mangifesta M, Ferrario C, Anzalone R, Viappiani A, Sinderen D van, Ventura M. 2017. Prevalence of antibiotic resistance genes among human gut-derived bifidobacteria. Applied and Environmental Microbiology. 83(3):e02894. doi:10.1128/AEM.02894-16

61. Xu F, Wang J, Guo Y, Fu P, Zeng H, Li Z, Pei X, Liu X, Wang S. 2018. Antibiotic resistance, biochemical typing, and PFGE typing of *Bifidobacterium* strains commonly used in probiotic health foods. Food Sci Biotechnol. 27(2):467–477. doi:10.1007/s10068-018-0320-6

62. Flórez AB, Ammor MS, Mayo B, van Hoek AHAM, Aarts HJM, Huys G. 2008. Antimicrobial susceptibility profiles of 32 type strains of *Lactobacillus*, *Bifidobacterium*, *Lactococcus* and *Streptococcus* spp. International Journal of Antimicrobial Agents. 31(5):484–486. doi:10.1016/j.ijantimicag.2007.09.003

63. Mayers DL, Sobel JD, Ouellette M, Kaye KS, Marchaim D, editors. 2017. Antimicrobial drug resistance: clinical and epidemiological aspects, Volume 2. Cham: Springer International Publishing. doi:10.1007/978-3-319-47266-9

64. Dai DLY, Petersen C, Hoskinson C, Del Bel KL, Becker AB, Moraes TJ, Mandhane PJ, Finlay BB, Simons E, Kozyrskyj AL, et al. 2023. Breastfeeding enrichment of *B. longum* subsp. *infantis* mitigates the effect of antibiotics on the microbiota and childhood asthma risk. Med. 4(2):92–112.e5. doi:10.1016/j.medj.2022.12.002

65. Filmer D, Pritchett LH. 2001. Estimating wealth effects without expenditure data-or tears: an application to educational enrollments in states of India. Demography. 38(1):115–132. doi:10.2307/3088292

