## Supplementary material for "Abundance of *Bifidobacterium* species in the infant gut microbiota and associations with maternal-infant characteristics in Dhaka, Bangladesh": Freitas_et_al_Bifidobacterium_Supplemental_material.zip

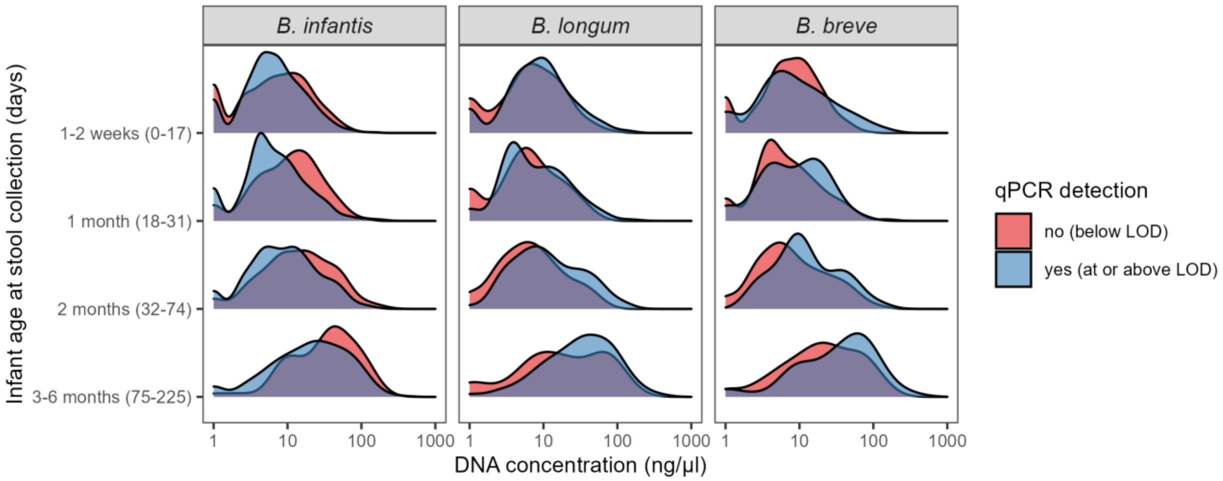


**Figure S1. DNA concentration distribution and qPCR detection status of *B. infantis, B. longum* and *B. breve*.**

Only samples tested for all three targets were included in this analysis (n=1120). The scale is fixed across all panels. LOD: limit of detection of the qPCR assay.

**A**

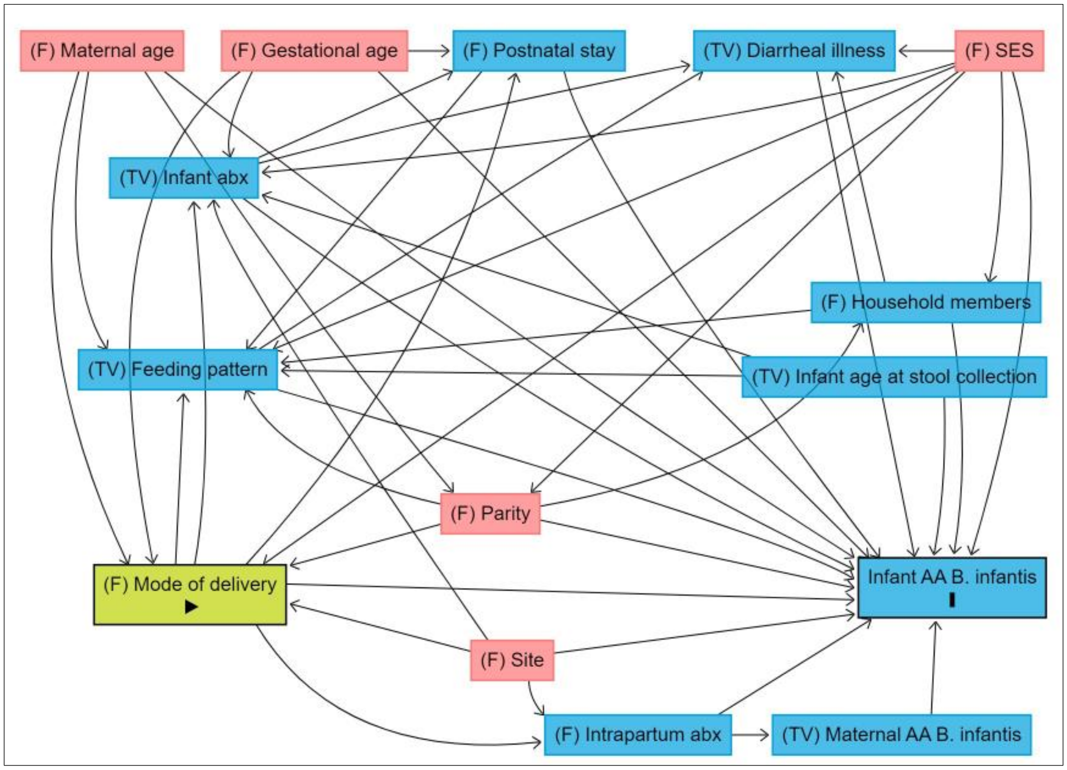


**B**

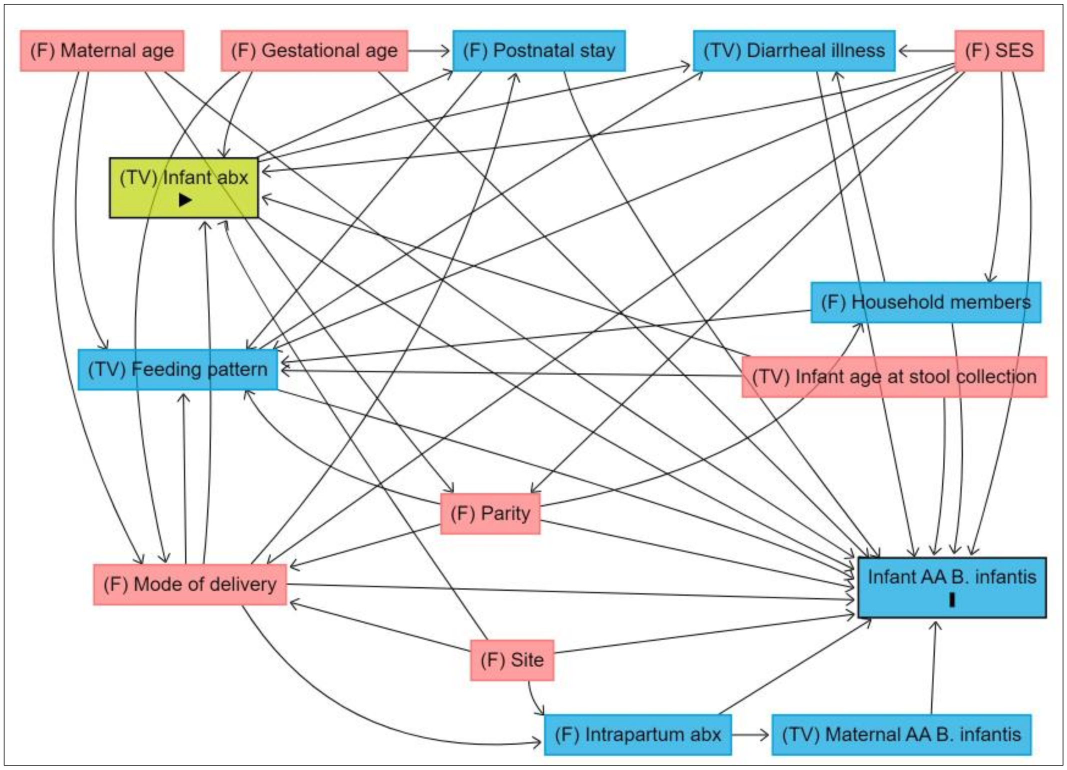


(Figure S2. continued on next page)

**C**

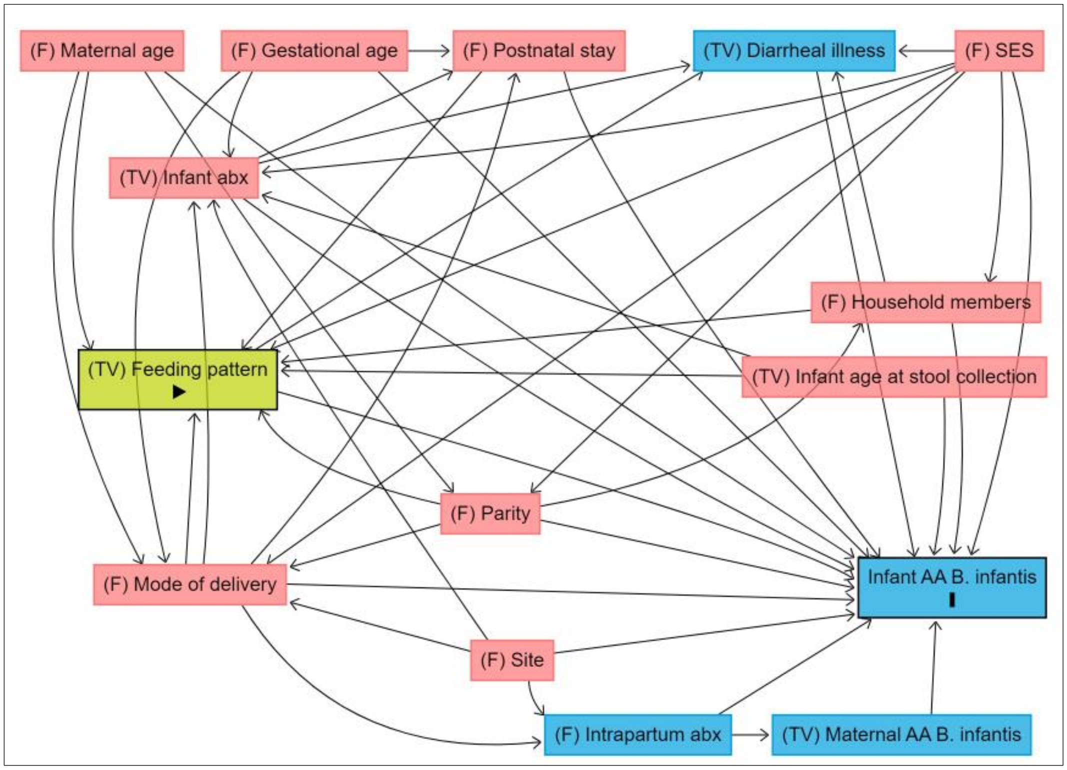


**Legend:**

- exposure
- outcome
- ancestor of exposure
- ancestor of outcome
- ancestor of exposure *and* outcome

**Figure S2. Directed Acyclic Graphs (DAG) of the association between *B. infantis* absolute abundance (AA) in infant stool samples and exposures.**

**(A)** Mode of delivery; **(B)** Infant antibiotic exposure; **(C)** Infant feeding pattern. TV: time varying variable; F: fixed variable; SES: socioeconomic status (represented by maternal education and asset index); abx: antibiotics.

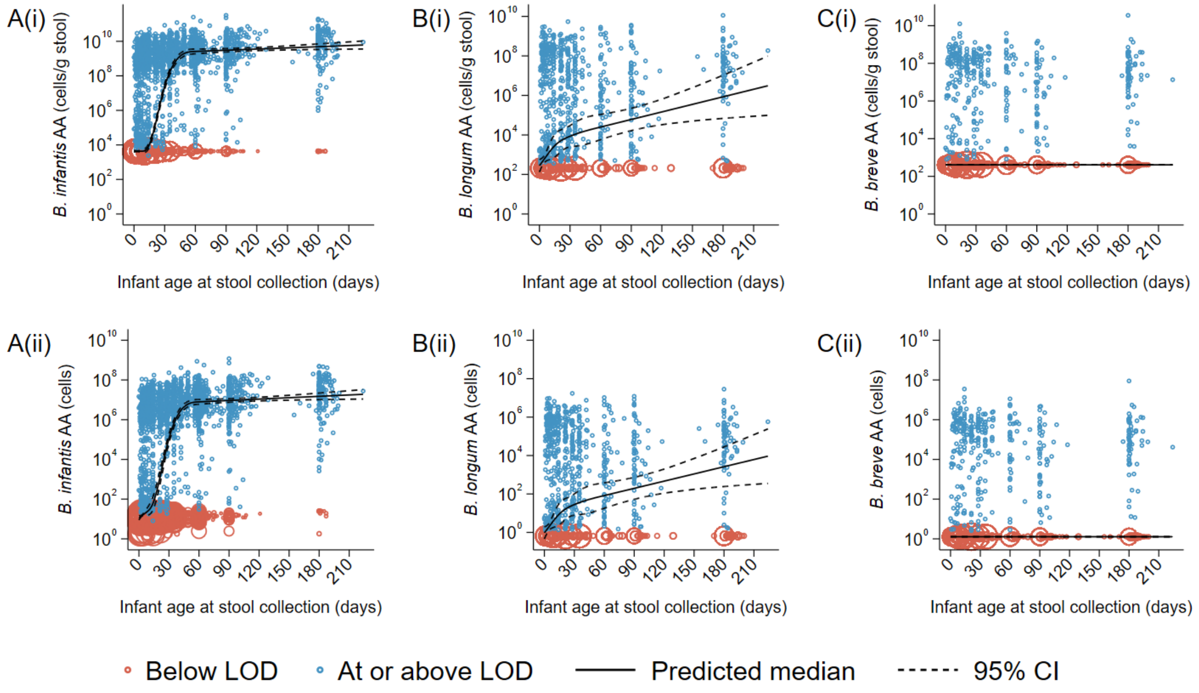


**Figure S3. Sensitivity analysis: *Bifidobacterium* (sub)species trajectory in early infancy, based on different normalization approaches: per stool mass (i) or not normalized (ii).**

**(A)** *B. infantis* (n=4613); **(B)** *B. longum* (n=1123); **(C)** *B. breve* (n=1161). Sample size (n) refers to the number of samples. The predicted median of absolute abundance (AA) and 95% confidence interval (95% CI) are based on a quantile regression model with clustered standard errors and restricted cubic splines (knots at 7, 14, 28 and 60 days of age). Samples below the assay limit of detection (LOD) were imputed as the median of one-half the LOD normalized to gram of stool, or not normalized. The size of each circle is proportional to the number of overlapping points.

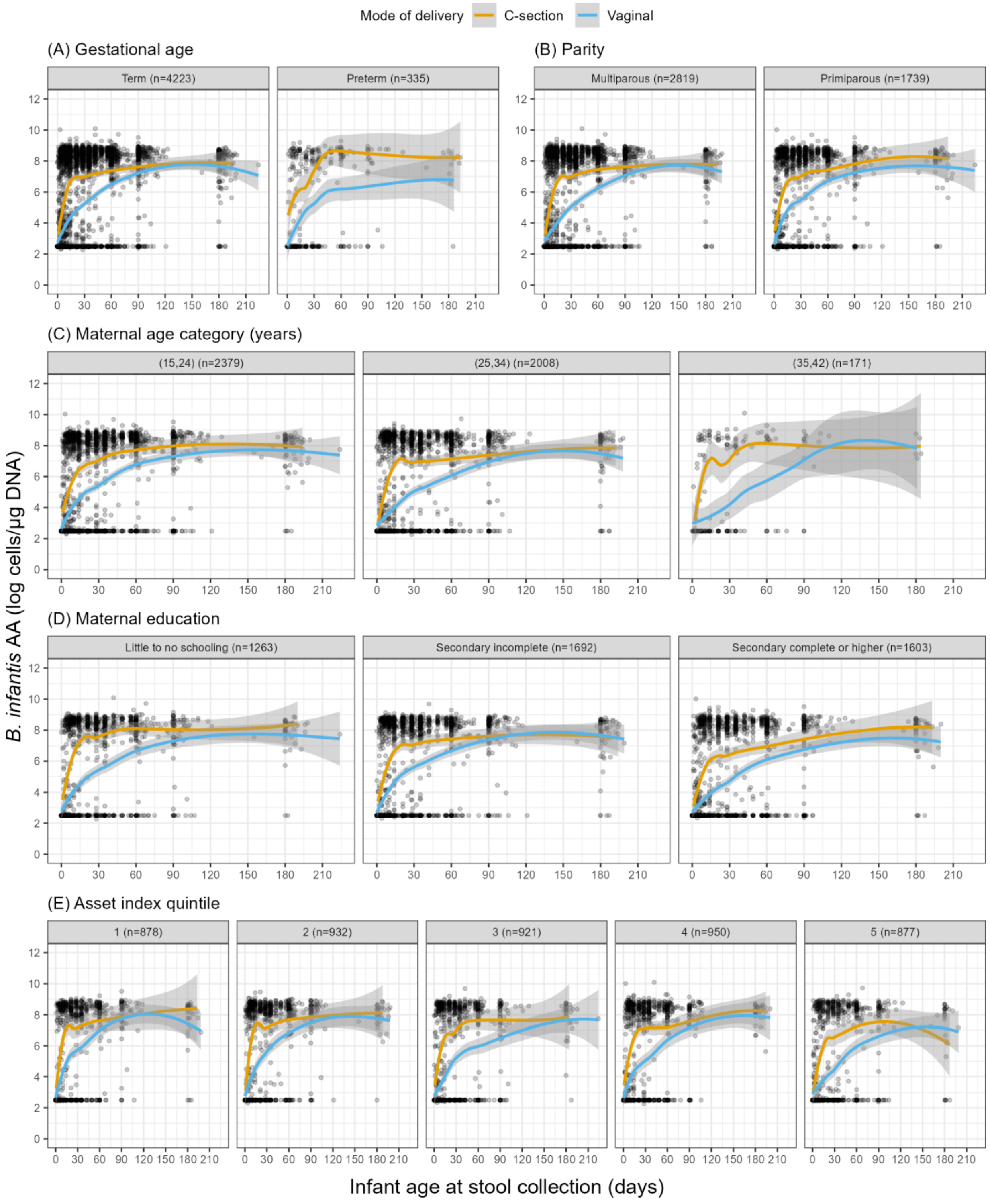


**Figure S4. *B. infantis* absolute abundance (AA) by mode of delivery, stratified by putative confounders.**

AA represented as loess curve (solid line) and 95% confidence interval (shade). Total samples, n=4558 (for all panels).

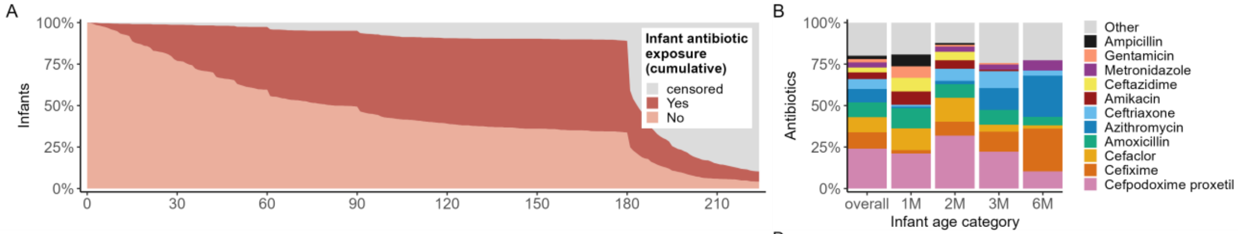


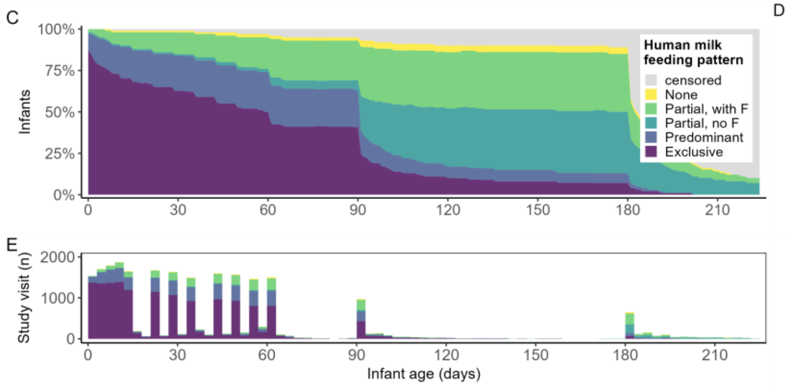


D

| Age [days] | At birth [0-4] | 1 month [5-34] | 2 months [35-76] | 3 months [77-174] | 6 months [175-224] |
|---|---|---|---|---|---|
| **Infants, n** | **181** | **277** | **361** | **296** | **778** |
| Formula | 31 (17) | 138 (50) | 211 (58) | 208 (70) | 387 (50) |
| Plain water | 29 (16) | 71 (26) | 141 (39) | 117 (40) | 508 (65) |
| Honey/dates | 134 (74) | 107 (39) | 116 (32) | 30 (10) | 71 (9) |
| Other liquids | 30 (17) | 26 (9) | 20 (6) | 19 (6) | 20 (3) |
| Solid foods | 2 (1) | 5 (2) | 14 (4) | 34 (11) | 632 (81) |

**Figure S5. Infant antibiotic exposure (A-B) and feeding pattern (C-E) up to six months of age (infants, n=1132).**

**(A)** Cumulative antibiotic exposure (‘Yes’ indicates previous exposure up to that day and ‘No’ indicates never being exposed). **(B)** Most common systemic antibiotics reported around 1M (0-34 days), 2M (35-76 days), 3M (77-174 days), and 6M (175-224 days) of age (M=month). Counts for antibiotics were determined at each calendar week (0-6, 7-13, 14-20, etc.), thus antibiotics reported in multiple visits within a week were counted only once (for example, if amoxicillin was reported at two scheduled visits on days 1 and 6, amoxicillin would be counted once at the first week of age); then counts were binned by age windows and the proportions calculated for plotting. The top 5 antibiotics in any age window were reported in addition to gentamicin and ampicillin (which are relevant for this population based on the WHO recommendation for management of infections in infants (0–59 days old) when referral to hospital is possible[1]. **(C)** Cumulative human milk feeding pattern from birth up until the next scheduled study visit. F=infant formula. **(D)** Most common foods reported among infants who did not consume human milk exclusively (number and percentage of infants who consumed each food category, n (%)) **(E)** Number of study visits for which feeding patterns were ascertained (bin width = 3 days). The artificial plateau in the feeding pattern along some intervals (e.g., 60-90 and 90-180 days) reflects the fewer study visits within those periods since most visits after two months of age happened around 60, 90 and 180 days of age. The ‘censored’ group in panels A/C corresponds to the proportion of infants from which there were no data collected due to loss to follow up (that is, infants did not complete all expected visits) or study exit (scheduled at day 180 but allowed to happen afterwards).

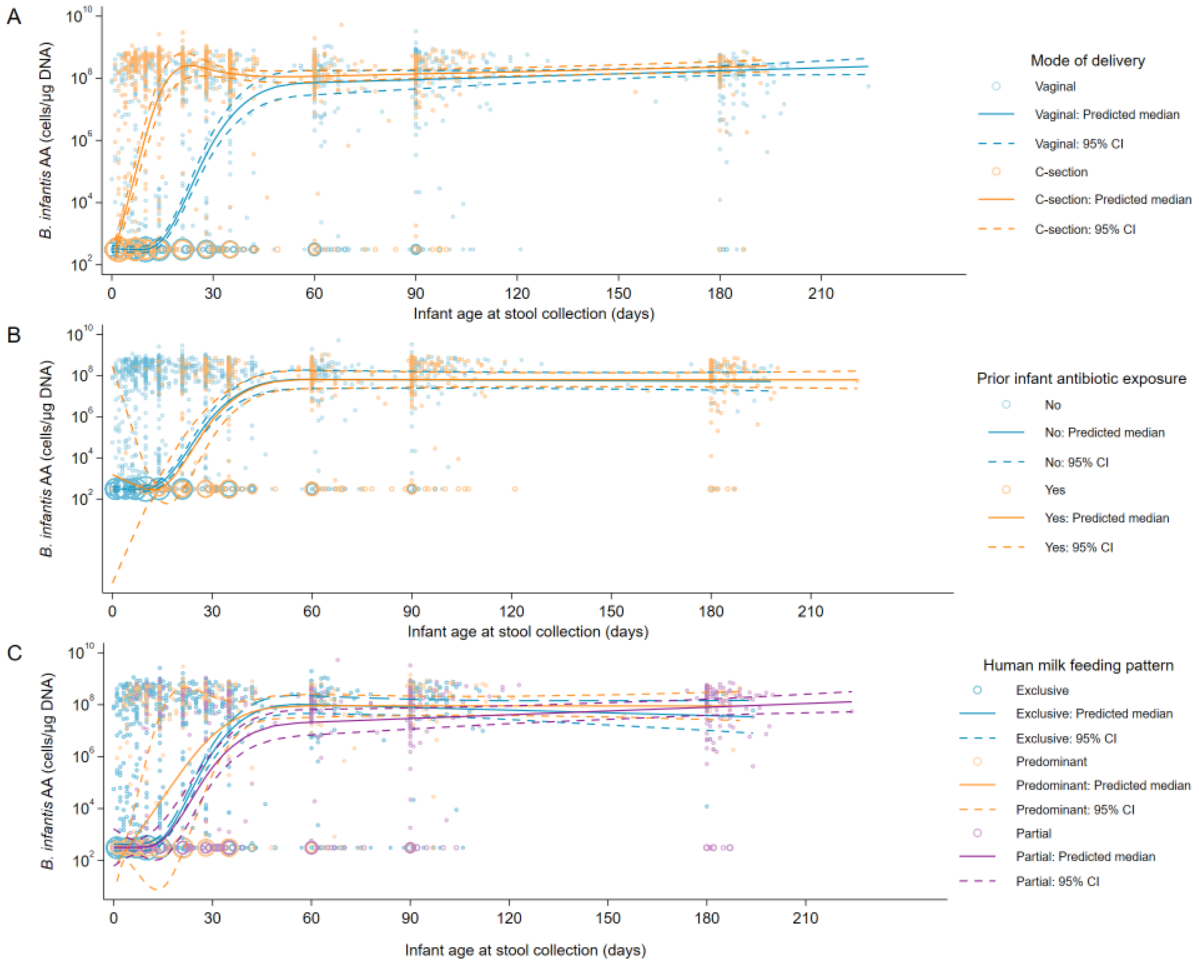


**Figure S6. Sensitivity analysis: *B. infantis* absolute abundance trajectory by infant-maternal characteristics, restricted to participants from schedules A and B.**

**(A)** Mode of delivery (n=3271); **(B)** Infant antibiotic exposure (n=3271); **(C)** Infant feeding pattern (n=3238). Sample size (n) refers to the number of samples. The predicted median of absolute abundance (AA) and 95% confidence interval (95% CI) are based on a quantile regression model with clustered standard errors and restricted cubic splines (knots at 7, 14, 28 and 60 days of age), adjusted for confounders. Samples below the assay limit of detection (LOD) were imputed as the median of one-half the LOD normalized to µg DNA. The size of each circle is proportional to the number of overlapping points. The 'none' category for infant feeding pattern was dropped due to small sample size (n=20 of 3238).

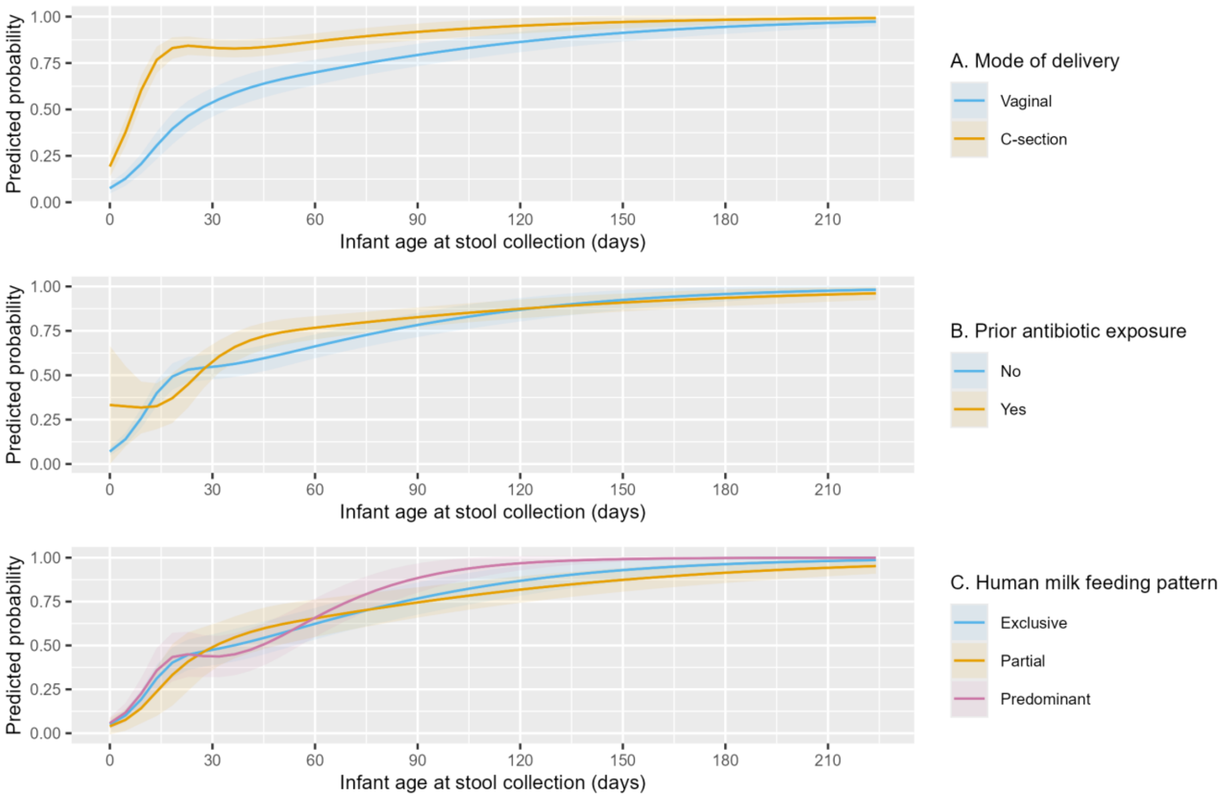


**Figure S7. Probability of high absolute abundance of *B. infantis*, by infant-maternal characteristics.** **(A)** Mode of delivery (n=4558); **(B)** Infant antibiotic exposure (n=4558); **(C)** Infant feeding pattern (n=4485). Sample size (n) refers to samples. The predicted probability of high *B. infantis* AA (≥6 log cells/µg DNA) (solid line) and 95% confidence interval (shade) are based on a Generalized Estimating Equations (GEE) model with restricted cubic splines (knots at 7, 14, 28 and 60 days of age), adjusted for confounders. The 'none' category for feeding pattern was dropped due to small sample size (n=36 of 4521).

**Table S1. qPCR primers and probes employed in this study.**

| Target | Name | Sequence (5' to 3') [a] | Length | Reference |
|---|---|---|---|---|
| *B. longum* subsp. *infantis* | inf_2348_F (Forward) | ATA CAG CAG AAC CTT GGC CT | 20 | Lawley et al 2017[2] |
| | inf_2348_R (Reverse) | GCG ATC ACA TGG ACG AGA AC | 20 | |
| | inf_2348_PB (Probe) | /6-FAM/ TTT CAC GGA /ZEN/ TCA CCG GAC CAT ACG /3IABkFQ/ | 24 | |
| | PhHV_F (Forward) | GGG CGA ATC ACA GAT TGA ATC | 21 | |
| | PhHV_R (Reverse) | GCG GTT CCA AAC GTA CCA A | 19 | |
| | PhHV_seq1 (probe) | 5'-Cy5 /TAT GTG TCC GCC ACC ATC T/3BHQ | 19 | |
| *B. longum* subsp. *longum* | lon_0274_F (Forward) | GAG GCG ATG GTC TGG AAG TT | 20 | Lawley et al 2017[2] |
| | lon_0274_R (Reverse) | CCA CAT CGC CGA GAA GAT TC | 20 | |
| | lon_0274_PB (Probe)[b] | /5TexRd-XN/ AAT TCG ATG CCC AGC GTG GTC TT /BHQ-2/ | 23 | |
| *B. breve* | bre_1623_F (Forward) | GTG GTG GCT TGA GAA CTG GAT AG | 23 | Haarman et al 2002[3] |
| | bre_1623_R (Reverse) | CAA AAC GAT CGA AAC AAA CAC TAA A | 25 | |
| | bre_1623_PB (Probe) | /6-FAM/ TGA TTC CTC GTT CTT GCT GT /MGB-NFQ/ | 20 | |

[a] *B. infantis, B. longum,* and *B. breve* primers were produced by ACGT Inc. (Toronto, ON, Canada); *B. infantis*, *B. longum*, and PhHV probes were produced by Integrated DNA technologies Canada Inc (IDT) (Toronto, ON, Canada); *B. breve* and PhHV primers were produced by Invitrogen Life Technologies™ (Burlington, ON, Canada).

[b] Probe modified from original design: 6-FAM reporter replaced with 5TexRd-XN, 3IABkFQ replaced with BHQ-2 and internal ZEN quencher removed.

Abbreviations: 6FAM = 5' 6-FAM™ reporter; ZEN = Internal ZEN™ quencher; IABkFQ = 3' Iowa Black® FQ quencher; Cy5 = Cy5 dye; 3BHQ = Black Hole quencher 3; 5TexRd-XN = 5' Texas Red-X (NHS Ester) reporter; BHQ-2 = 3' Black Hole Quencher-2; MGB-NFQ = Minor Groove Binder – 3' Nonfluorescent Quencher.

**Table S2. Infant, maternal, and household characteristics, by enrollment study site.**

| | MCHTI | MFSTC | *p-value* [k] |
|---|---|---|---|
| ***Delivery and infant characteristics: infants, n (%)*** [a] | **452 (40)** | **680 (60)** | |
| Infant age at enrollment (days), median ($25^{th}$, $75^{th}$)<br>[min, max] | 1 (1, 2)<br>[0, 4] | 1 (0, 1)<br>[0, 4] | <0.001 |
| Gestational age at delivery (weeks), median ($25^{th}$, $75^{th}$) | 39.3 (38.3, 40.1) | 39.1 (38.1, 40.1) | 0.5 |
| Term (≥ 37 weeks), n (%) | 429 (95) | 612 (91) | 0.01 |
| Preterm (< 37 weeks), n (%) | 23 (5) | 62 (9) | |
| Mode of delivery, n (%) | | | <0.001 |
| Vaginal | 171 (38) | 351 (52) | |
| C-section | 281 (62) | 329 (48) | |
| Birth weight [b] (g), median ($25^{th}$, $75^{th}$) | 2840 (2580, 3060) | 2860 (2600, 3150) | 0.2 |
| Sex: male infants, n (%) | 208 (46) | 331 (49) | 0.4 |
| Singletons, n (%) | 444 (98) | 671 (99) | 0.7 |
| Postnatal hospital stay (days), median ($25^{th}$, $75^{th}$) | 4 (1, 5) | 2 (1, 4) | <0.001 |
| Vaginal delivery | 1 (1, 1) | 1 (0, 1) | <0.001 |
| C-section delivery | 5 (4, 5) | 4 (4, 4) | <0.001 |
| Infant antibiotic exposure [c], n (%) | | | |
| Up to 1 month of age | 125 (28) | 100 (15) | <0.001 |
| Up to 2 months of age | 233 (53) | 222 (33) | <0.001 |
| Up to 6 months of age | 329 (77) | 300 (51) | <0.001 |
| Human milk feeding pattern [d], n (%) | Exclusive \| Predominant<br>Partial \| None | Exclusive \| Predominant<br>Partial \| None | |
| At or near enrollment | 386 (86) \| 55 (12)<br>10 (2.2) \| 0 (0) | 602 (89) \| 56 (8.3)<br>16 (2.4) \| 2 (0.3) | 0.1 |
| At 1 month of age | 241 (59) \| 99 (24)<br>67 (16) \| 4 (1.0) | 437 (69) \| 118 (19)<br>69 (11) \| 7 (1.1) | 0.004 |
| At 2 months of age | 190 (43) \| 116 (26)<br>123 (28) \| 11 (2.5) | 386 (58) \| 150 (23)<br>117 (18) \| 11 (1.7) | <0.001 |
| At 6 months of age | 25 (6.1) \| 45 (11)<br>314 (76) \| 28 (6.8) | 54 (11) \| 25 (5.1)<br>400 (82) \| 9 (1.8) | <0.001 |
| Diarrheal illness [e], n (%) | | | |
| Up to 1 month of age | 31 (7.0) | 12 (1.8) | <0.001 |
| Up to 2 months of age | 59 (13) | 29 (4.3) | <0.001 |
| Up to 6 months of age | 126 (29) | 69 (12) | <0.001 |
| ***Mother and household characteristics, n (%)*** | **448 (40)** | **677 (60)** | |
| Maternal age (years), median ($25^{th}$, $75^{th}$) | 24 (20, 27) | 24 (20, 28) | 0.7 |
| Parity, median ($25^{th}$, $75^{th}$) | 2 (1, 2) | 2 (1, 2) | >0.9 |
| Primiparous, n (%) | 185 (41) | 269 (40) | 0.6 |
| Maternal peripartum antibiotics [f], n (%) | | | <0.001 |
| None | 24 (5.4) | 33 (4.9) | |
| Intrapartum only | 16 (3.6) | 49 (7.2) | |
| Postpartum only | 256 (57) | 548 (81) | |
| Both intrapartum and postpartum | 152 (34) | 47 (6.9) | |
| Maternal education, n (%) | | | <0.001 |
| Little to no schooling [g] | 95 (21) | 223 (33) | |
| Secondary incomplete | 173 (39) | 206 (30) | |
| Secondary complete or higher | 180 (40) | 248 (37) | |
| Asset index quintile [h], n (%) | | | 0.002 |
| 1 (lowest wealth) | 64 (14) | 142 (21) | |
| 2 | 90 (20) | 134 (20) | |
| 3 | 82 (18) | 145 (22) | |
| 4 | 119 (27) | 124 (18) | |
| 5 (highest wealth) | 89 (20) | 126 (19) | |
| Household size [i], median ($25^{th}$, $75^{th}$) | 5 (4, 7) | 5 (4, 6) | <0.001 |
| One or more household members ≤ 5 years old [j], n (%) | 157 (37) | 196 (31) | 0.05 |

[a] Missing data: gestational age (n=6, 0.5%); infant antibiotic exposure (n=13 (1.1%), n=22 (1.9%), n=117 (10%) for 1, 2 and 6 months of age, respectively); human milk feeding pattern (n=5 (0.4%), n=90 (8%), n=28 (2.5%), n=232 (20%), for 1, 2 and 6 months of age, respectively); diarrheal illness (n=13 (1.1%), n=23 (2%), n=116 (10%) for 1, 2 and 6 months of age, respectively); asset index (n=10, 0.9%); household size (n=17, 1.5%); members ≤ 5 years old (n=58, 5.2%).

[b] Weight measured by study personnel was used as a birthweight proxy for four infants where there was >15% difference between birthweight and weight measured within 1-4 days of birth; for three infants, weight was measured within 1 day of birth and, for one infant, weight measured within 4 days of birth was used.

[c] Antibiotic exposure status was derived as any exposure versus no exposure within age windows around the scheduled visits at 1 month (25-31 days), 2 months (46-74 days) and 6 months (173-225 days). Data were considered missing for an infant if their last study visit happened before the age window cut-off.

[d] Human milk feeding pattern was derived based on the infant's feeding status at the closest scheduled visit available at enrollment (0-4 days), and at 1, 2 and 6 months (same age windows as defined in 'c'). Data were considered missing for an infant if there were no study visits within the age window.

[e] Diarrheal illness status was defined as any previous episode of diarrhea (versus never), reported at scheduled visits around 1, 2 and 6 months of age (same age windows and criteria for missing data as defined in 'c').

[f] Intrapartum refers to any antibiotics administered before labour (at hospital admission in which delivery occurred), during labour and/or in the operating theatre, but prior to delivery. Postpartum refers to antibiotics administered after the mother has delivered the infant.

[g] Includes women with no formal education, and incomplete and completed primary school.

[h] Asset index scores were generated using Principal Components Analysis (PCA) for all participants enrolled in the observational cohort study (n=1886) and in a concurrently running trial (n=519) at the same study sites with the same eligibility criteria. The score is a summary measure of household wealth based on ownership of the following assets: electricity, radio, television, almirah, fan, table, chair, fridge, pump, freezer, phone, animals, mobile, watch, computer, autobike, bicycle, rickshaw and a vehicle[4].

[i] Number of people that sleep in the same house as the infant enrolled in the study, including the infant's twin, if applicable, but excluding the infant enrolled in the study.

[j] Excludes the infant enrolled in the study.

[k] P-values: Wilcoxon rank-sum tests were used to compare medians of continuous variables, which were all non-normally distributed; Chi-square tests were used to compare categorical variables when expected counts were ≥ 5. Fisher's exact tests were used to compare categorical variables when expected counts were < 5.

**Table S3. Sample size for qPCR analysis, by enrollment schedule (A, B, C).**

| | | ***B. infantis*** | | | | ***B. longum*** | | | | ***B. breve*** | | | |
|---|---|---|---|---|---|---|---|---|---|---|---|---|---|
| **Scheduled visit (days since birth)** | **Infant age in days (min-max)** | **Total** | **A** | **B** | **C** | **Total** | **A** | **B** | **C** | **Total** | **A** | **B** | **C** |
| 0-1 | 0-2 | 421 | 51 | 226 | 144 | 63 | 4 | 58 | 1 | 69 | 4 | 58 | 7 |
| 3-4 | 3-5 | 486 | 55 | 231 | 200 | 80 | 12 | 67 | 1 | 94 | 12 | 67 | 15 |
| 6-7 | 6-8 | 476 | 71 | 273 | 132 | 110 | 22 | 88 | 0 | 122 | 22 | 88 | 12 |
| 10 | 9-10 | 394 | 66 | 216 | 112 | 73 | 19 | 54 | 0 | 77 | 19 | 54 | 4 |
| 14 | 11-17 | 453 | 76 | 304 | 73 | 113 | 19 | 94 | 0 | 114 | 19 | 94 | 1 |
| 21 | 18-24 | 398 | 68 | 254 | 76 | 119 | 28 | 91 | 0 | 118 | 28 | 90 | 0 |
| 28 | 25-31 | 413 | 65 | 250 | 98 | 116 | 26 | 90 | 0 | 119 | 26 | 93 | 0 |
| 35 | 32-45 | 486 | 70 | 261 | 155 | 115 | 22 | 93 | 0 | 113 | 22 | 91 | 0 |
| 60 | 46-74 | 597 | 58 | 245 | 294 | 100 | 20 | 80 | 0 | 101 | 20 | 81 | 0 |
| 90 | 75-130 | 324 | 64 | 244 | 16 | 108 | 26 | 82 | 0 | 108 | 26 | 82 | 0 |
| 180 | 131-225 | 165 | 41 | 124 | 0 | 126 | 34 | 92 | 0 | 126 | 34 | 92 | 0 |
| **Total infant samples** | - | **4613** | 685 | 2628 | 1300 | **1123** | 232 | 889 | 2 | **1161** | 232 | 890 | 39 |
| **Total infants** | - | **1132** | 75 | 287 | 770 | **364** | 75 | 287 | 2 | **399** | 75 | 287 | 37 |
| **Total mother** [a] | 0-7 | **491** | 66 | 164 | 261 | - | - | - | - | - | - | - | - |

[a] Sample size refers to 491 mothers with 495 infants (i.e., 4 twin pairs), where each mother provided one sample within the specified age window.

**Table S4. Absolute abundance of *B. infantis*, *B. longum* and *B. breve* at discrete infant ages.** [a]

| Target | Age (days) | Predicted median: Log cells per µg DNA (95% CI) | Predicted median: Log cells per gram stool (95% CI) | Predicted median: Log cells (95% CI) |
|---|---|---|---|---|
| *B. infantis* | 0 | 2.49 (2.45, 2.54) | 3.63 (3.58, 3.69) | 1.06 (0.96, 1.16) |
| | 7 | 2.49 (2.45, 2.54) | 3.63 (3.58, 3.69) | 1.32 (1.18, 1.46) |
| | 14 | 2.76 (2.64, 2.87) | 3.89 (3.75, 4.02) | 1.78 (1.45, 2.12) |
| | 21 | 4.14 (4.01, 4.26) | 5.21 (5.05, 5.38) | 3.08 (2.74, 3.43) |
| | 28 | 5.94 (5.83, 6.06) | 6.96 (6.79, 7.12) | 4.67 (4.42, 4.92) |
| | 35 | 7.25 (7.14, 7.35) | 8.22 (8.06, 8.38) | 5.81 (5.63, 6.00) |
| | 42 | 7.98 (7.88, 8.08) | 8.95 (8.79, 9.11) | 6.47 (6.32, 6.63) |
| | 60 | 8.38 (8.30, 8.47) | 9.41 (9.28, 9.55) | 6.90 (6.78, 7.02) |
| | 90 | 8.30 (8.23, 8.37) | 9.48 (9.38, 9.58) | 6.97 (6.88, 7.06) |
| | 180 | 8.06 (7.93, 8.19) | 9.69 (9.53, 9.85) | 7.18 (7.01, 7.34) |
| *B. longum* | 0 | 1.24 (0.90, 1.58) | 2.44 (2.13, 2.76) | -0.03 (-0.34, 0.29) |
| | 7 | 1.82 (1.56, 2.09) | 2.94 (2.69, 3.19) | 0.46 (0.21, 0.70) |
| | 14 | 2.36 (1.78, 2.95) | 3.40 (2.85, 3.96) | 0.91 (0.37, 1.45) |
| | 21 | 2.70 (2.12, 3.29) | 3.71 (3.15, 4.26) | 1.21 (0.66, 1.76) |
| | 28 | 2.89 (2.37, 3.42) | 3.89 (3.37, 4.41) | 1.40 (0.87, 1.93) |
| | 35 | 3.02 (2.43, 3.61) | 4.03 (3.43, 4.64) | 1.55 (0.92, 2.18) |
| | 42 | 3.11 (2.47, 3.75) | 4.15 (3.48, 4.82) | 1.67 (0.98, 2.37) |
| | 60 | 3.25 (2.65, 3.86) | 4.40 (3.76, 5.04) | 1.92 (1.26, 2.58) |
| | 90 | 3.44 (2.83, 4.05) | 4.78 (4.22, 5.35) | 2.29 (1.72, 2.86) |
| | 180 | 4.01 (2.53, 5.48) | 5.93 (4.84, 7.02) | 3.42 (2.39, 4.45) |
| *B. breve* [b] | 0 | 1.51 | 2.61 | 0.11 |
| | 7 | 1.51 | 2.61 | 0.11 |
| | 14 | 1.51 | 2.61 | 0.11 |
| | 21 | 1.51 | 2.61 | 0.11 |
| | 28 | 1.51 | 2.61 | 0.11 |
| | 35 | 1.51 | 2.61 | 0.11 |
| | 42 | 1.51 | 2.61 | 0.11 |
| | 60 | 1.51 | 2.61 | 0.11 |
| | 90 | 1.51 | 2.61 | 0.11 |
| | 180 | 1.51 | 2.61 | 0.11 |

[a] Predicted median of absolute abundance (AA) and 95% confidence interval (95% CI) are based on a quantile regression model with clustered standard errors and restricted cubic splines (knots at 7, 14, 28 and 60 days of age). AA was treated as the dependent continuous variable, and infant age was the primary independent continuous variable. Samples with values below the assay limit of detection (LOD) were imputed as a single value represented by the median of half the LOD (from samples for which LOD imputation was applied) normalized to µg DNA, gram of stool or not normalized. Number of samples: *B. infantis* (n=4613); *B. longum* (n=1123); *B. breve* (n=1161).

[b] The predicted median AA in all ages was equivalent to the imputed value since the majority of data points were below LOD, therefore, given the little variation in AA, measures of precision could not be estimated.

**Table S5. Absolute abundance of *B. infantis* at discrete infant ages, by mode of delivery, infant antibiotic exposure and infant feeding pattern. [a]**

| Mode of delivery (samples, n=4558) | | | |
|---|---|---|---|
| | Predicted median: log cells per µg DNA (95% CI) | | |
| Age (days) | Vaginal (reference) | C-section | Difference between groups |
| 0 | 2.51 (2.47, 2.55) | 1.64 (1.54, 1.74) | -0.86 (-0.97, -0.75) |
| 7 | 2.49 (2.46, 2.52) | 4.69 (4.50, 4.87) | 2.20 (2.02, 2.39) |
| 14 | 2.70 (2.63, 2.78) | 7.37 (7.08, 7.66) | 4.67 (4.37, 4.97) |
| 21 | 3.95 (3.80, 4.09) | 8.44 (8.19, 8.69) | 4.50 (4.21, 4.78) |
| 28 | 5.58 (5.36, 5.81) | 8.46 (8.33, 8.59) | 2.88 (2.62, 3.14) |
| 35 | 6.78 (6.49, 7.06) | 8.35 (8.27, 8.43) | 1.58 (1.29, 1.87) |
| 42 | 7.46 (7.16, 7.77) | 8.29 (8.20, 8.38) | 0.83 (0.51, 1.15) |
| 60 | 7.91 (7.62, 8.20) | 8.27 (8.18, 8.36) | 0.36 (0.06, 0.66) |
| 90 | 7.99 (7.78, 8.20) | 8.29 (8.22, 8.36) | 0.30 (0.08, 0.53) |
| 180 | 8.23 (8.10, 8.35) | 8.35 (8.20, 8.51) | 0.13 (-0.07, 0.33) |

| Infant antibiotic exposure (samples, n=4558) | | | |
|---|---|---|---|
| | Predicted median: log cells per µg DNA (95% CI) | | |
| Age (days) | No exposure (reference) | Prior exposure | Difference between groups |
| 0 | 2.67 (2.46, 2.87) | 3.08 (1.65, 4.52) | 0.50 (-0.93, 1.93) |
| 7 | 3.01 (2.74, 3.28) | 2.86 (2.35, 3.37) | -0.06 (-0.58, 0.45) |
| 14 | 3.52 (3.03, 4.02) | 2.91 (2.39, 3.43) | -0.53 (-1.15, 0.10) |
| 21 | 4.76 (4.30, 5.22) | 4.11 (3.45, 4.77) | -0.57 (-1.32, 0.18) |
| 28 | 6.23 (5.93, 6.53) | 5.78 (5.32, 6.24) | -0.37 (-0.88, 0.14) |
| 35 | 7.27 (7.05, 7.50) | 6.99 (6.69, 7.30) | -0.20 (-0.52, 0.13) |
| 42 | 7.86 (7.65, 8.08) | 7.69 (7.44, 7.93) | -0.09 (-0.33, 0.14) |
| 60 | 8.19 (7.98, 8.40) | 8.10 (7.88, 8.32) | -0.01 (-0.19, 0.18) |
| 90 | 8.14 (7.96, 8.31) | 8.10 (7.92, 8.27) | 0.04 (-0.11, 0.20) |
| 180 | 7.97 (7.74, 8.21) | 8.10 (7.89, 8.30) | 0.20 (-0.11, 0.52) |

| Infant feeding pattern (samples, n=4485) | | | | | |
|---|---|---|---|---|---|
| | Predicted median: log cells per µg DNA (95% CI) | | | | |
| Age (days) | Exclusive human milk (reference) | Predominant human milk | Partial human milk | Difference between Predominant & Exclusive | Difference between Partial & Exclusive |
| 0 | 2.68 (2.52, 2.83) | 1.82 (1.06, 2.57) | 2.39 (1.03, 3.75) | -0.92 (-1.66, -0.18) | -0.29 (-1.63, 1.05) |
| 7 | 2.89 (2.71, 3.07) | 3.77 (2.48, 5.06) | 3.00 (2.33, 3.68) | 0.82 (-0.50, 2.14) | 0.11 (-0.57, 0.79) |
| 14 | 3.31 (3.01, 3.61) | 5.58 (2.71, 8.45) | 3.72 (1.51, 5.93) | 2.21 (-0.69, 5.11) | 0.41 (-1.82, 2.64) |
| 21 | 4.59 (4.31, 4.88) | 6.75 (4.07, 9.43) | 4.89 (2.68, 7.10) | 2.09 (-0.61, 4.79) | 0.29 (-1.94, 2.51) |
| 28 | 6.18 (5.96, 6.40) | 7.39 (5.86, 8.93) | 6.14 (4.80, 7.47) | 1.15 (-0.40, 2.70) | -0.04 (-1.39, 1.30) |
| 35 | 7.31 (7.11, 7.51) | 7.79 (7.14, 8.44) | 7.02 (6.38, 7.66) | 0.42 (-0.24, 1.08) | -0.29 (-0.94, 0.37) |
| 42 | 7.94 (7.74, 8.14) | 8.02 (7.78, 8.26) | 7.53 (7.24, 7.83) | 0.02 (-0.24, 0.28) | -0.41 (-0.73, -0.09) |
| 60 | 8.27 (8.08, 8.45) | 8.20 (7.97, 8.44) | 7.87 (7.65, 8.08) | -0.13 (-0.38, 0.12) | -0.40 (-0.62, -0.18) |
| 90 | 8.15 (8.00, 8.31) | 8.30 (8.12, 8.47) | 7.93 (7.76, 8.11) | 0.08 (-0.12, 0.28) | -0.22 (-0.41, -0.03) |
| 180 | 7.81 (7.47, 8.16) | 8.57 (7.99, 9.15) | 8.13 (7.93, 8.32) | 0.69 (0.02, 1.37) | 0.31 (-0.08, 0.70) |

[a] Predicted median of absolute abundance (AA) and 95% confidence interval (95% CI) are based on a quantile regression model with clustered standard errors and restricted cubic splines (knots at 7, 14, 28 and 60 days of age), adjusted for confounders [b]. AA was treated as the dependent continuous variable and exposure was included as a main effect and in an interaction term with chronologic age of the infants. Samples with values below the assay limit of detection (LOD) were imputed as a single value represented by the median of half the LOD normalized to μg DNA. The 'none' category for feeding pattern was dropped due to small sample size (n=36 of 4521).

[b] Each model was adjusted for the following covariates: *Mode of delivery*: Study site (binary), Gestational age at birth (continuous), Maternal age (continuous), Parity (binary), Maternal education (categorical), Asset index (continuous); all variables are time-fixed. *Infant antibiotic exposure*: Study site (binary), Gestational age at birth (continuous), Mode of delivery (binary), Maternal education (categorical), Asset index (continuous); all variables are time-fixed. *Infant feeding pattern*: Study site (binary), Gestational age at birth (continuous), Mode of delivery (binary), Postnatal hospital stay (continuous), Maternal age (continuous), Parity (binary), Maternal education (categorical), Asset index (continuous), Household size (continuous); all variables are time-fixed.

**Table S6. Candidate mediators (M) of the association between mode of delivery (MOD) and *B. infantis* absolute abundance (AA).** [a]

| Candidate mediator | Age (days) | Samples (n) | Infants (n) | Model description and Coefficient (95% CI) [b] | | | |
|---|---|---|---|---|---|---|---|
| | | | | AA ~ MOD<br>Coefficient of MOD | AA ~ MOD + M<br>Coefficient of MOD | AA ~ M<br>Coefficient of M | M ~ MOD<br>Coefficient of MOD [c] |
| Length of postnatal hospital stay (days) [d] | 3-17 | 1700 | 743 | 5.38 (4.84, 5.93) | 5.38 (4.85, 5.92) | 0.86 (0.47, 1.25) | 4.66 (4.15, 5.22) |
| | 3-31 | 2495 | 851 | 5.61 (5.42, 5.80) | 5.61 (5.38, 5.84) | 1.18 (1.03, 1.34) | 4.74 (4.25, 5.29) |
| | 3-45 | 2997 | 939 | 5.65 (5.52, 5.78) | 5.65 (5.46, 5.84) | 1.25 (1.14, 1.36) | 4.82 (4.33, 5.36) |
| | 3-17 | 743 [e] | 743 | 4.89 (3.89, 5.89) | 4.89 (3.46, 6.32) | 0.79 (0.47, 1.11) | 4.66 (4.15, 5.22) |
| | 3-31 | 851 [e] | 851 | 5.51 (5.22, 5.80) | 5.51 (5.10, 5.92) | 1.01 (0.76, 1.25) | 4.74 (4.25, 5.29) |
| Maternal intrapartum antibiotics (yes) | 3-17 | 1809 | 792 | 4.71 (3.72, 5.69) | 4.71 (3.72, 5.69) | 0.64 (-0.79, 2.07) | 11.55 (6.99, 19.09) |
| | 3-31 | 2604 | 893 | 5.40 (5.08, 5.72) | 5.40 (5.08, 5.73) | 2.22 (-2.14, 6.58) | 11.59 (7.19, 18.67) |
| | 3-45 | 3106 | 973 | 5.54 (5.35, 5.74) | 5.54 (5.35, 5.74) | 3.91 (1.30, 6.53) | 10.95 (6.94, 17.27) |

[a] All models were adjusted for study site and were restricted to samples collected at age windows within approximately the first month of age (samples collected at days 0-2 were dropped since they had low AA variability due to overall low detection rate). An interaction term with age (restricted cubic splines) was not added to facilitate interpretation of the coefficients. In all models involving MOD, the reference group was 'Vaginal delivery'. All models with *B. infantis* AA as outcome were based on a quantile regression model. Rows within the same 'candidate mediator' panel represent models restricted to different age windows.

[b] Model description: outcome ~ exposure, where the + sign indicates addition of candidate mediator into the model (if applicable). Example: AA ~ MOD + M (6th column in the table) refers to model assessing the effect of mode of delivery on *B. infantis* AA, adjusted for the candidate mediator (as listed in the first column).

[c] Models with postnatal hospital stay as outcome were based on a Poisson regression, and the coefficients are shown as incidence rate ratio. Models with intrapartum antibiotic as outcome were based on a logistic regression, and the coefficients are shown as odds ratio.

[d] Analysis excluded samples collected prior to hospital discharge.

[e] Included only the first post-discharge sample.

**Table S7. Association of maternal stool *B. infantis* detection (in the first week postpartum) and infant *B. infantis* absolute abundance (AA) during the first month of age.** [a]

| **Variable** | **Difference in infant *B. infantis* AA Regression coefficient (95% CI)** | | | |
|---|---|---|---|---|
| | **Unadjusted** | **Adjusted for delivery mode** | **Adjusted for study site** | **Adjusted for delivery mode and study site** |
| Maternal *B. infantis* detection (above LOD) | 5.60 (5.11, 6.08) | 0.49 (-0.17, 1.16) | 5.60 (5.11, 6.08) | 0.49 (-0.17, 1.16) |
| C-section | - | 5.28 (4.74, 5.82) | - | 5.28 (4.73, 5.83) |
| Study site (MFSTC) | - | - | 0.00 (-0.043, 0.043) | 0.00 (-0.095, 0.095) |

[a] All coefficients were based on a quantile regression model with clustered standard errors. Maternal stool *B. infantis* was treated as a time-fixed, independent binary (detected vs not-detected) variable based on the qPCR result of the first maternal stool sample available within 0-7 days postpartum. Infant *B. infantis* AA was treated as the dependent, continuous variable. Analysis was restricted to infant samples collected in the first month of age (3-31 days) and included only infant samples whose mothers also had a sample collected. Infant samples collected prior to their mother sample were not included in the analysis (n=1481 samples, 429 infants). LOD: denotes limit of detection.

**Supplemental references**


1. WHO. 2015. (World Health Organization) Guideline: managing possible serious bacterial infection in young infants when referral is not feasible. Geneva: World Health Organization. https://iris.who.int/handle/10665/181426

2. Lawley B, Munro K, Hughes A, Hodgkinson AJ, Prosser CG, Lowry D, Zhou SJ, Makrides M, Gibson RA, Lay C, et al. 2017. Differentiation of *Bifidobacterium longum* subspecies *longum* and *infantis* by quantitative PCR using functional gene targets. PeerJ. 5:e3375. doi:10.7717/peerj.3375

3. Haarman M, Knol J. 2005. Quantitative real-time PCR assays to identify and quantify fecal *Bifidobacterium* species in infants receiving a prebiotic infant formula. Applied and Environmental Microbiology. 71(5):2318–2324. doi:10.1128/AEM.71.5.2318-2324.2005

4. Filmer D, Pritchett LH. 2001. Estimating wealth effects without expenditure data-or tears: an application to educational enrollments in states of India. Demography. 38(1):115–132. doi:10.2307/3088292
